# Frontoparietal network integrity supports cognitive function despite atrophy and hypoperfusion in pre-symptomatic frontotemporal dementia: multimodal analysis of brain function, structure and perfusion

**DOI:** 10.1101/2024.03.06.24303617

**Authors:** Xulin Liu, P Simon Jones, Maurice Pasternak, Mario Masellis, Arabella Bouzigues, Lucy L Russell, Phoebe H. Foster, Eve Ferry-Bolder, John van Swieten, Lize Jiskoot, Harro Seelaar, Raquel Sanchez-Valle, Robert Laforce, Caroline Graff, Daniela Galimberti, Rik Vandenberghe, Alexandre de Mendonça, Pietro Tiraboschi, Isabel Santana, Alexander Gerhard, Johannes Levin, Sandro Sorbi, Markus Otto, Florence Pasquier, Simon Ducharme, Chris Butler, Isabelle Le Ber, Elizabeth Finger, Maria Carmela Tartaglia, Matthis Synofzik, Fermin Moreno, Barbara Borroni, Jonathan D. Rohrer, Kamen A. Tsvetanov, James B. Rowe, The GENFI consortium

## Abstract

**INTRODUCTION:** Genetic mutation carriers of frontotemporal dementia can remain cognitively well despite neurodegeneration. A better understanding of brain structural, perfusion and functional patterns in pre-symptomatic stage could inform accurate staging and potential mechanisms.

**METHODS:** We included 207 pre-symptomatic genetic mutation carriers and 188 relatives without mutations. The grey matter volume, cerebral perfusion, and resting-state functional network maps were co-analyzed using linked independent component analysis (LICA). Multiple regression analysis was used to investigate the relationship of LICA components to genetic status and cognition.

**RESULTS:** Pre-symptomatic mutation carriers showed an age-related decrease in the left frontoparietal network integrity while non-carriers did not. Executive functions of mutation carriers became dependent on the left frontoparietal network integrity in older age.

**DISCUSSION:** The frontoparietal network integrity of pre-symptomatic mutation carriers showed a distinctive relationship to age and cognition compared to non-carriers, suggesting a contribution of the network integrity to brain resilience, despite atrophy and hypoperfusion.

## 1. Background

Frontotemporal dementia (FTD) is characterized by the selective degeneration of the frontal and temporal cortices, leading to progressive deficits in behavior, social and executive function, or language [1]. Genetic risk factors are important, with about 20-30% of FTD cases being familial [2]. Highly penetrant mutations in three major genes, chromosome 9 open reading frame 72 (*C9orf72*), microtubule-associated protein tau (*MAPT*), and progranulin (*GRN*), account for about 60% of cases of familial FTD [1]. Given that neurobiological changes could occur many years before the onset of symptoms of neurodegenerative dementias [3-6], investigation at the early stage of diseases before symptom onset is important for understanding factors that facilitate the brain’s resilience. Genetic FTD with highly penetrant genetic mutations provides the opportunity for early investigation before symptom onset. Comparison between pre-symptomatic genetic mutation carriers and their family members without the mutation, allows one to investigate the effect of early neurodegeneration without the confounding influence of medication and lifestyle changes after symptom onset.

People carrying highly penetrant genetic mutations of FTD have grey matter atrophy and reduction in cerebral blood flow (CBF) more than a decade before the expected symptom onset, as measured by magnetic resonance imaging (MRI) and arterial spin labelling (ASL) [4, 6-8]. However, functional network organization and connectivity are generally maintained despite significant atrophy in pre-symptomatic genetic FTD [4, 9]. Moreover, a recent study indicates that functional networks predict cognitive decline and symptomatic conversion in pre-symptomatic genetic mutation carriers [10]. A better understanding of these changes in the pre-symptomatic stage would inform accurate staging, facilitate clinical trials, and elucidate the mechanisms of resilience by which gene carriers remain cognitively well for many years despite biomarker evidence of neurodegeneration.

Here we test whether pre-symptomatic differences in brain structure, cerebral perfusion and functional network act synergistically or independently on clinically relevant disease features such as cognitive performance and disease progression. Specifically, we used linked independent components analysis of multimodal imaging to investigate whether the interplay of brain grey matter atrophy, cerebral perfusion and functional network integrity explains difference between pre-symptomatic FTD genetic mutation carriers and non-carriers.

## 2. Methods

### 2.1 Participants

The Genetic Frontotemporal dementia Initiative (GENFI) study is an international muti-centre cohort study across Europe and Canada. GENFI recruited participants with genetic mutations of FTD and their relatives [6, 7]. Participants included carriers of genetic mutations in *C9orf72, GRN,* and *MAPT* who have or have not shown symptoms, and their relatives without genetic mutations. Most participants are unaware of their genetic status at recruitment, and remain unaware of their genetic status by a genetic-guardianship process. Participants underwent a standardized clinical assessment consisting of a medical history, family history, and physical examination. Symptomatic status was based on the assessment by clinicians to determine whether the participants fulfilled the diagnostic criteria for FTD [11-13]. Functional status was measured using the Frontotemporal Dementia Rating Scale [14] and behavioral symptoms were assessed using the Cambridge Behavioural Inventory Revised version (CBI-R) [15]. We also tested the Mini-Mental State Examination (MMSE). Participants not diagnosed with FTD who had functional, cerebrovascular and structural neuroimaging data with good quality were included in this study (N = 395). There were 207 FTD genetic mutation carriers who had not shown symptoms fulfilling the diagnostic criteria for FTD at the time of recruitment, termed pre-symptomatic genetic mutation carriers. The pre-symptomatic genetic mutation carriers had no symptoms, and were not manifesting abnormal behaviours that next of kin reported to be out of the normal range. There were 188 relatives of the mutation carriers who are not genetic mutation carriers of known FTD genes, termed non-carriers. The demographics of the subjects are shown in **Table 1**. Demographic variables were compared between pre-symptomatic mutation carriers and non-carriers using one-way Analysis of Variance (ANOVA) for continuous variables and using the Chi-squared test for categorical variables.

**Table 1.**
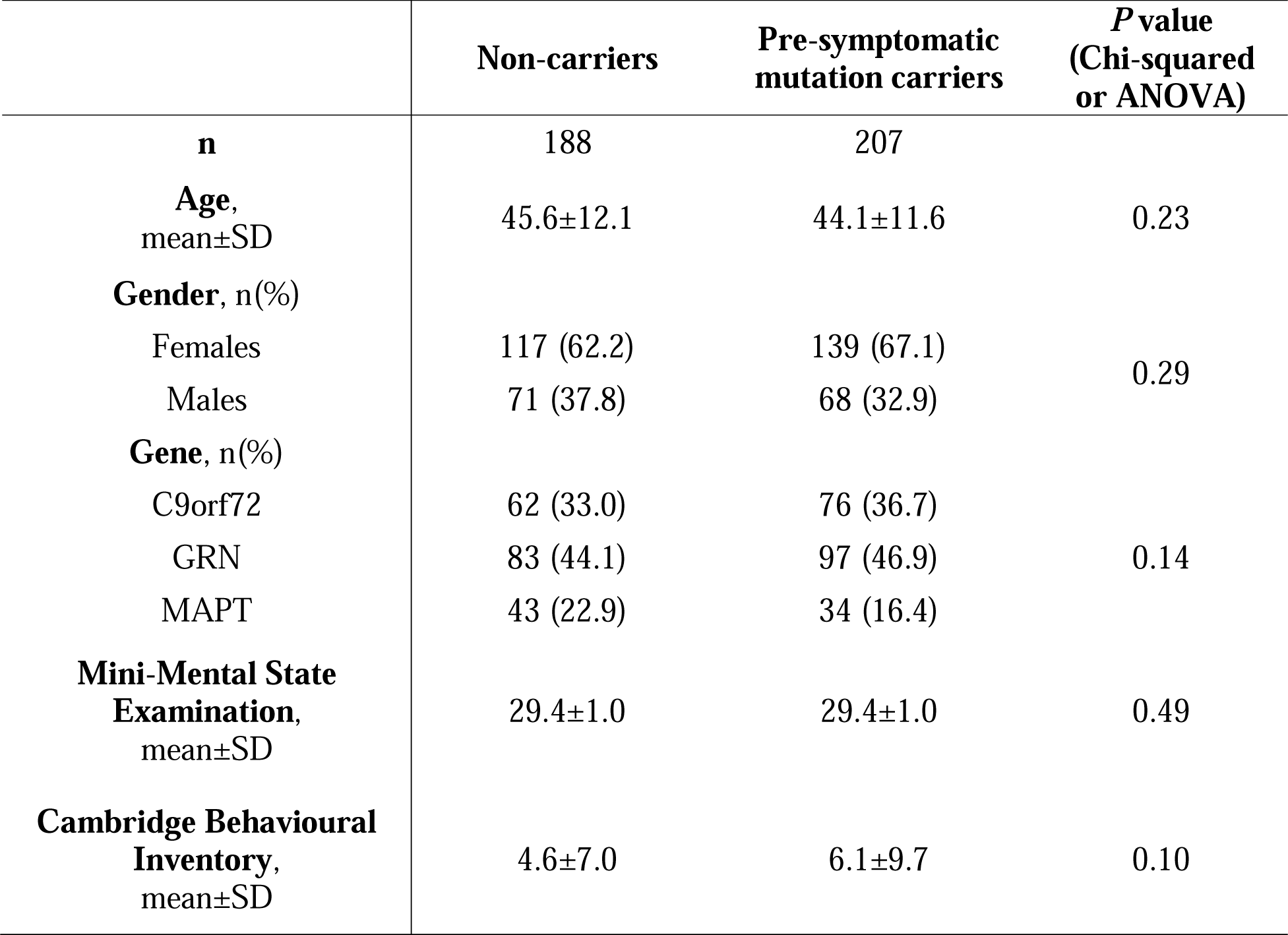
Characteristics of participants.

|  | **Non-carriers** | **Pre-symptomatic**<br>**mutation carriers** | ***P* value**<br>**(Chi-squared**<br>**or ANOVA)** |
| --- | --- | --- | --- |
| **n** | 188 | 207 |  |
| **Age,**<br>mean±SD | 45.6±12.1 | 44.1±11.6 | 0.23 |
| **Gender, n(%)** |  |  |  |
| Females | 117 (62.2) | 139 (67.1) | 0.29 |
| Males | 71 (37.8) | 68 (32.9) |  |
| **Gene, n(%)** |  |  |  |
| C9orf72 | 62 (33.0) | 76 (36.7) | 0.14 |
| GRN | 83 (44.1) | 97 (46.9) |  |
| MAPT | 43 (22.9) | 34 (16.4) |  |
| **Mini-Mental State**<br>**Examination,**<br>mean±SD | 29.4±1.0 | 29.4±1.0 | 0.49 |
| **Cambridge Behavioural**<br>**Inventory,**<br>mean±SD | 4.6±7.0 | 6.1±9.7 | 0.10 |

### 2.2 Neurocognitive assessment

Participants underwent a neuropsychological battery consisting of tests from the Uniform Data Set [16], covering attention and processing speed: Wechsler Memory Scale–Revised (WMS-R) digit span forward [16], Trail-Making Test part A (TMTA) [17], the Wechsler Adult Intelligence Scale-Revised (WAIS-R) Digit Symbol Substitution test [16], Delis-Kaplan Executive Function System (DKEFS) Color-Word Interference Test color and word naming [18]; executive function: WMS-R Digit span backward [16], TMT part B (TMTB) [17], DKEFS Color-Word Interference Test ink naming [18]; language: modified Camel and Cactus Test [19], the Boston Naming Test (short 30-item version) [16], verbal fluency: category fluency and phonemic fluency [16, 20]; memory encoding: Free and Cued Selective Reminding Test (FCSRT) immediate free and total recall [21]; memory recall: FCSRT delayed free and total recall, Benson Complex Figure recall [21]; and visuoconstruction: Benson Complex Figure copy. More details of the neurocognitive assessment in this cohort can also be found in the previously published protocol [6]. A principal component analysis (PCA) with permutation (n = 1000) was performed on the series of cognitive measures. Leading components were selected for further investigation.

### 2.3 Neuroimaging acquisition and processing

#### 2.3.1 Grey matter volume

T1-weighted MRI scans were collected on 3T scanners. A three-dimensional T1-weighted magnetization prepared rapid gradient echo sequence image was acquired for each subject accommodating different scanners at each site over at least 283 s (283 to 462 s) and had a median isotropic resolution of 1.1 mm (1 to 1.3 mm), repetition time (TR) of 2000 ms (6.6 to 2400), echo time (TE) of 2.9 ms (2.6 to 3.5 ms), inversion time of 8 ms (8 to 9 ms), and field of view (FOV) 256 × 256 × 208 mm (192 to 256 × 192 to 256 × 192 to 208 mm). For participants with baseline and follow-up scans, the latest available scans were examined. The co-registered T1 images were segmented to extract probabilistic maps of six tissue classes: grey matter, white matter, cerebrospinal fluid, bone, soft tissue, and residual noise. The native-space grey matter and white matter images were submitted to diffeomorphic registration to create equally represented gene-group template images [22]. The templates for all tissue types were normalized to the Montreal Neurological Institute (MNI) template using a 12-parameter affine transformation. The normalized and modulated grey matter volume (GMV) images were used in analysis.

#### 2.3.2 CBF

ASL sequences could be different across different sites. The sequences included in this study were: pseudo-continuous ASL 3D fast-spin-echo stack-of-spirals implemented on a 3T General Electric MR750; pseudo-continuous ASL 2D gradient-echo echo-planar imaging on a 3T Philips Achieva, with and without background suppression; and pulsed ASL 3D gradient-and-spin-echo on 3T Siemens Trio systems. The complete ASL parameters of each sequence have been described elsewhere [23].

For ASL processing, the ExploreASL pipeline (v1.5.1) was used [24]. The ExploreASL is optimized for multi-center data through the use of advanced ASL markers (e.g., spatial coefficient-of-variation [25] and partial volume correction [26]). It has been employed so far in over 30 studies, consisting of ASL scans from three MRI vendors including GE, Philips, and Siemens [24]. A recent study using this ASL processing method to analyze cerebral perfusion data from the GENFI study has also confirmed the reliability of this method for integrating ASL data from different scanners specific to the GENFI cohort data [27]. This denoising for scanner effects was complemented with data-driven and model-driven correction at the subject level [28, 29]. In this study, structural and functional image volumes across multiple sites, vendors, and sequences were processed first. Briefly, structural images were non-linearly registered to MNI space using Geodesic Shooting [30] and transformation matrices were saved for subsequent application on functional images. ASL scans were corrected for motion outliers using rigid-body transformation coupled with the enhancement of automated blood flow estimates outlier exclusion algorithm [31], followed by pairwise subtraction to produce perfusion-weighted images. Functional proton-density weighted images were smoothed with a 16mm full width at half maximum (FWHM) Gaussian kernel to create a bias field that avoided division artifacts during CBF quantification and cancelled out acquisition-specific B1-field inhomogeneities. CBF quantification itself followed a single-compartment model approach and recommendations outlined in the ASL consensus paper [32]. For quality control, CBF images were reviewed independently by three authors with 3-6 years of experience in handling ASL data. Disagreements were resolved by consensus. CBF volumes were masked by their structural T1 counterpart’s probability grey matter mask at ≥ 50% and the spatial coefficient of variation was calculated for the extracted voxels. Images with coefficient of variation values ≥ 0.8 were discarded.

To adjust for differences arising from the effects of multiple sites, scanners, and software, a spatially varying intensity normalization approach was used [8], together with data-driven and model-driven approaches at the between-subject level (see section Statistical analysis). The normalization approach uses the within-site CBF similarity between participants to remove the between-site quantification differences [8]. Mean CBF images of these groupings were calculated and smoothed using a 6.4mm FWHM Gaussian kernel. Smoothing was constrained to a binary MNI brain mask. These group-specific mean images were then averaged to calculate the population mean CBF image, which in turn was rescaled uniformly such that the mean grey matter perfusion equalled 60 ml / min / 100 g. Group-specific rescale-factor images were then calculated by dividing this population CBF image by each group’s mean CBF image. Individual CBF images were adjusted via multiplication against their group’s respective rescale-factor image. To account for the effects of atrophy, partial volume correction on rescaled CBF volumes was performed using a linear regression approach [26]. Further details of ASL processing are discussed in a recent publication [27]. Due to hyperintensities present in the cerebellum of many subjects which is not our interest of study, only the CBF of the cortical region was included in the analysis of this study. A cortical binary mask created from the Harvard-Oxford cortical atlas (https://fsl.fmrib.ox.ac.uk/fsl/fslwiki/Atlases) was therefore applied to all CBF images.

#### 2.3.3 Resting-state functional networks

For rs-fMRI, echo planar imaging acquired 200 volumes with 42 slices (slice thickness = 3.5 mm, TR = 2500 ms; TE = 30 ms; FOV = 192 mm x 192 mm). Resting-state fMRI data were preprocessed using Automatic Analysis [33] calling functions from SPM12 implemented in Matlab (MathWorks). Processing steps included (1) spatial realignment to correct for head movement and movement by distortion interactions, (2) temporal realignment of all slices, and (3) coregistration of the echo planar imaging to the participant’s T1 anatomical scan. The normalization parameters from the T1 stream were applied to warp functional images into MNI space. Resting-state fMRI data were further processed using whole-brain ICA of single-subject time series denoising, with noise components selected and removed automatically using the ICA-based Automatic Removal of Motion Artifacts toolbox [34]. This was complemented with linear detrending of the fMRI signal, covarying out six realignment parameters, white matter and cerebrospinal fluid signals, their first derivatives, and quadratic terms [35]. Global white matter and cerebrospinal fluid signals were estimated for each volume from the mean value of white matter and cerebrospinal fluid masks derived by thresholding SPM tissue probability maps at 0.75. Data were band-pass filtered using a discrete cosine transform.

In order to identify the activation of functional networks from rs-fMRI, group-level ICA was performed to decompose the rs-fMRI data (<u>trendscenter.org/software/gift/</u>) [36] from 395 participants (including pre-symptomatic mutation carriers and non-carriers). ICA dissociates signals from complex datasets with minimal assumptions, to represent data in a small number of independent components (ICs) which here are spatial maps that describe the temporal and spatial characteristics of underlying signals [36, 37]. The values at each voxel reflect the correlation between the time series of the voxel and that of the component. Each component can therefore be interpreted as BOLD co-activation across voxels of a functional network at resting state [38]. The number of components used, N = 15, matched a common degree of decomposition previously applied in low-dimensional group-ICA of rs-fMRI [39-41] and generated network spatial maps that showed a high degree of overlapping with network templates. Low-dimensional group-ICA was used because the purpose was to define each network with a single component, and high-dimensional group-ICA would tend to decompose single network into multiple components. The stability of the estimated ICs was evaluated across 100 ICASSO iterations [42]. Functional networks were identified from components by visualization and validated by spatially matching the components to pre-existing templates [43], in accordance with the previous methodology used to identify networks from ICs [44]. The dorsal and ventral default mode network, the salience network, and the left and right frontoparietal network were selected, which are higher-order functional networks known to be associated with age- and FTD-related cognitive change [45-47].

### 2.4 Statistical analysis

#### 2.4.1 Linked ICA

Linked independent component analysis (ICA) was performed using FLICA of FMRIB (https://fsl.fmrib.ox.ac.uk/fsl/fslwiki/FLICA) [48, 49] implemented in Matlab (MathWorks version 2021b). Linked ICA is a data-driven analytic method that allows for simultaneous characterization of multimodal imaging modalities while taking into account the covariance across imaging modalities [48]. In comparison with other commonly used multivariate approaches for multivariate data integration such as canonical correlation analysis and partial least squares, linked ICA is able to identify patterns of covariance across more than two modalities. Linked ICA was run with seven spatial map inputs: GMV, CBF, and five co-activation maps from resting-state functional networks (i.e., the dorsal default mode network, the ventral default mode network, the salience network, the right frontoparietal network and the left frontoparietal network) identified as described in 2.3.3. To ensure the results were not influenced dominantly by non-grey matter regions (e.g., ventricles), all spatial maps were masked by thresholding SPM grey matter tissue probability maps at 0.3. We refer to these imaging-derived spatial maps as modalities in linked ICA. A summary flow chart of the processing and analysis of imaging modalities is presented in **Figure 1**.

**Figure 1.**
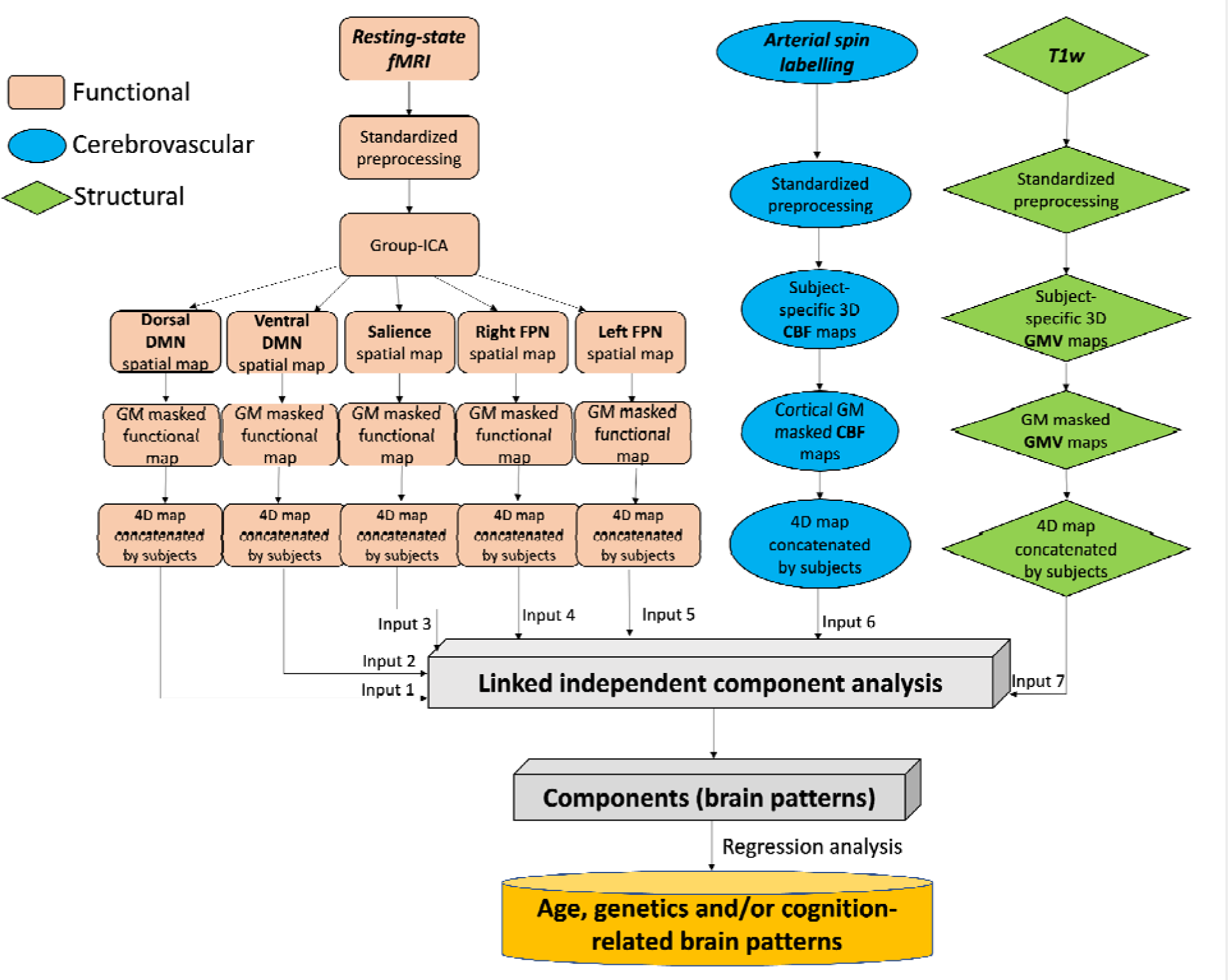
Summary of processing and analysis of the imaging modalities, comprising functional, cerebrovascular, and structural measurements. CBF, cerebral blood flow; DMN, default mode network; FPN, frontoparietal network; GMV, grey matter volume; ICA, independent component analysis; fMRI, functional magnetic resonance imaging; SN, salience network; T1w, T1-weighted.

Within each modality, images from all subjects were concatenated into a single input matrix (participants-by-voxels) for linked ICA. Linked ICA decomposed this n-by-m matrix of participants-by-voxels into spatial components, with each component being an aggregate of spatial patterns, one for each modality, along with a set of subject loadings, one for each component [48]. Each modality spatial pattern is a map of weights that is later converted to pseudo-Z-statistic by accounting for the scaling of the variables and the signal-to-noise ratio in that modality. Only modalities with significant contribution (i.e., weighting with Z-score > 3.34, which corresponds to *P* < 0.001) were presented in this study. Linked ICA subject loadings for a given component were shared among all modalities represented in that component and indicated the degree to which that component was expressed by any individual subject. Subject loadings were used as inputs to the second-level between-subject regression analysis (see below in 2.4.2).

#### 2.4.2 Multiple regression analysis

To investigate the effects of age (linear and quadratic) and genetic mutation on cognition, multiple regression analysis was used with cognition PCA component scores as the dependent variable. Group was classified by genetic mutation status (i.e., pre-symptomatic mutation carriers or non-carriers). Gender and site effect were included as covariates. In Wilkinson’s notation [50], the model took the form:

*Cognition component ∼ Group*Age^2 + Gender + site*.

To investigate whether brain patterns were predicted by age (linear and quadratic), genetic mutation and their interaction, subject loadings of each linked ICA component (IC) of interest were investigated as the dependent variable in multiple regression. Gender, total brain volume and site effect were included as covariates. In Wilkinson’s notation, the model took the form:

*IC ∼ Group*Age^2 + Gender + total brain volume + site*.

Finally, to investigate the relationship between brain patterns and cognitive variability, accounting for the effects of genetics and age (linear and quadratic), multiple regression was used taking the following form:

*Cognition component ∼ IC*Group*Age^2 + Gender + total brain volume + site*.

A false discovery rate (FDR)-corrected *P* < 0.05 was considered statistically significant. Analyses were performed in Matlab.

## 3. Results

### 3.1 Relationship between age, gene group and cognitive function

The two significant PCA components are shown in **Figure 2**. The first cognition component (variance explained 36.6%, *P* < 0.001) was related to global cognitive function. No significant group-wise difference in global cognition was found between genetic mutation carriers and non-carriers (*P* = 0.079). Both non-carriers and pre-symptomatic genetic mutation carriers showed a decline in global cognition with age likely reflecting the general age-related decrease in global cognitive function. No significant difference was found in the age-cognition relationship between genetic mutation carriers and non-carriers (Group:Age interaction t = -0.97, *P* = 0.33; Group:Age^2 interaction t = -0.73, *P* = 0.47).

**Figure 2.**
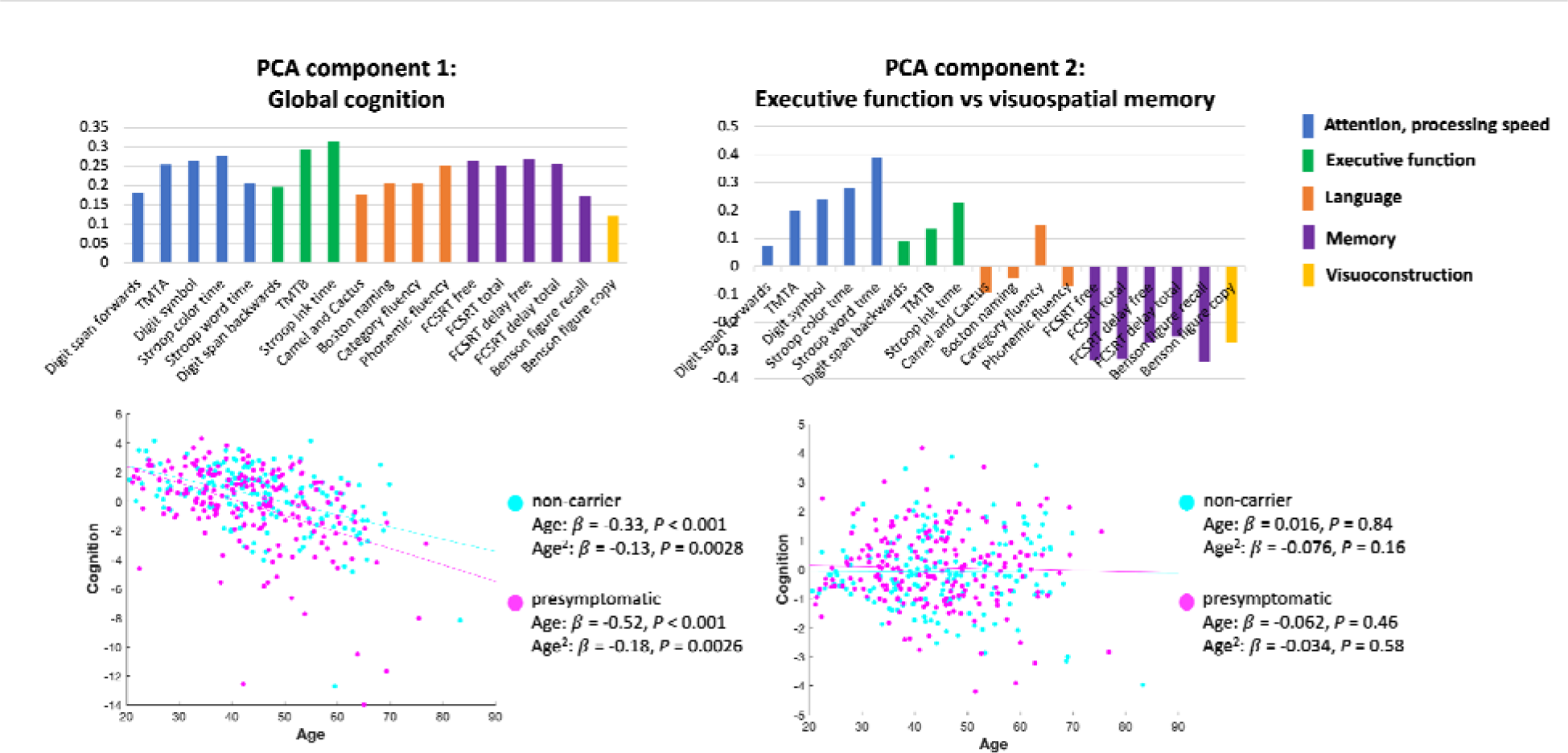
The two significant components from principal component analysis (PCA) on cognitive assessments. The top row shows the loadings of each cognitive test in PCA components. The bottom row shows the scatter plots of the correlation between age and PCA cognition component scores.

The second cognition component (variance explained 9.1%, *P* < 0.001) indicated executive function, attention and processing speed with deficits in visuospatial memory. No significant group-wise difference was found between genetic mutation carriers and non-carriers (*P* = 0.28). Neither non-carriers nor pre-symptomatic genetic mutation carriers showed age-related change in these cognitive functions. No significant difference was found in the age-cognition relationship between genetic mutation carriers and non-carriers (Group:Age interaction t = -0.62, *P* = 0.53; Group:Age^2 interaction t = 0.58, *P* = 0.56).

### 3.2 Multimodal fusion using linked ICA

The relative weight of modalities in each linked ICA output component is shown in **Figure 3**. Three components (IC10, IC14 and IC19) were excluded from further analysis as they were dominated by signals from one or two subjects (e.g., due to regional hyperintensities reflected by ASL images). We focused on components with variance explained > 1%. Note that there was little fusion between functional signals and structural or vascular signals.

**Figure 3.**
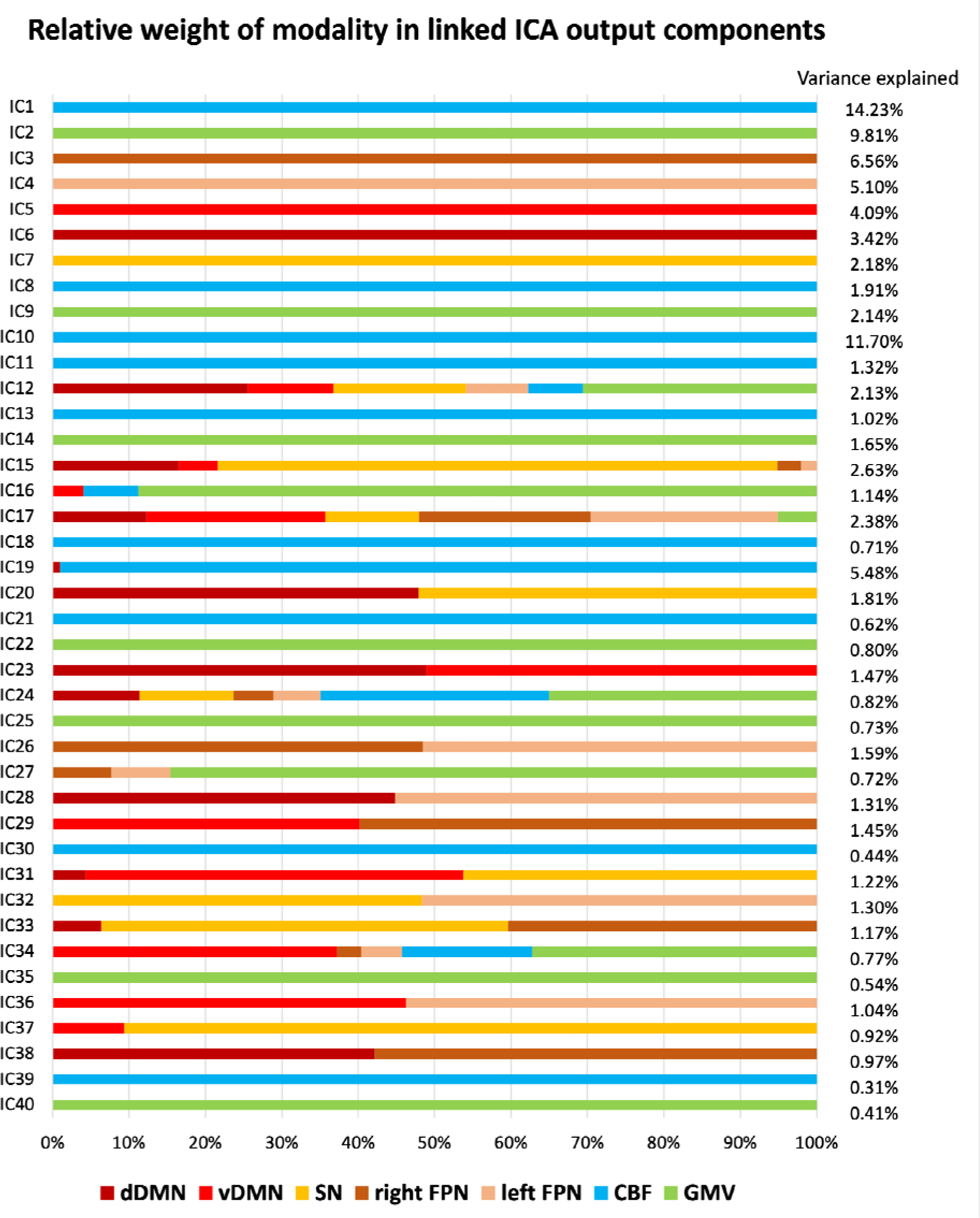
The relative weight of modalities in each component generated from linked independent component analysis (ICA) and the percentage of variance explained of each component. CBF, cerebral blood flow; dDMN, dorsal default mode network; vDMN, ventral default mode network; FPN, frontoparietal network; GMV, grey matter volume; SN, salience network.

### 3.3 Relationship between age, gene group and neuroimaging components

Multiple regression analysis results of the linked ICA components of interest are shown in **Table 2**. We focused on components with a significant model fit (FDR-corrected *P* < 0.05 for adjusted R^2^, i.e., the components that showed significant correlations with the variables being tested). Strong linear age effects were observed particularly in components indicating global CBF (IC1), ventral default mode network (IC5), salience network (IC7), and head motion (IC9) (**Figure 4**). Only one component, IC4, showed differential age effects between pre-symptomatic and non-carriers (Group:Age interaction t = -2.82, *P* = 0.0051). As age increases, pre-symptomatic genetic mutation carriers showed decreased activation of the left frontoparietal network (IC4, r = -0.30, *P* < 0.001), while non-carriers did not (r = -0.0087, *P* = 0.91). Brain visualization of IC4 and its scatter plot against age are shown in **Figure 5**. Further analyses to examine for possible specificity to *GRN*, *MAPT* or *C9orf72* carriers showed that the interaction between genetic mutation status and age (Group:Age) in the regression model was significant within the *GRN* mutation carriers (Group:Age interaction t = -2.44, *P* = 0.016), but was not significant in the rest of the pre-symptomatic genetic mutation carriers excluding *GRN* mutation carriers (Group:Age interaction t = -1.43, *P* = 0.16). It was neither significant within the *C9orf72* mutation carriers (Group:Age interaction t = -1.53, *P* = 0.13) nor within the *MAPT* mutation carriers (Group:Age interaction t = -1.42, *P* = 0.16) alone. Brain spatial maps of other components are presented in **Supplementary** Figure 1.

**Figure 4.**
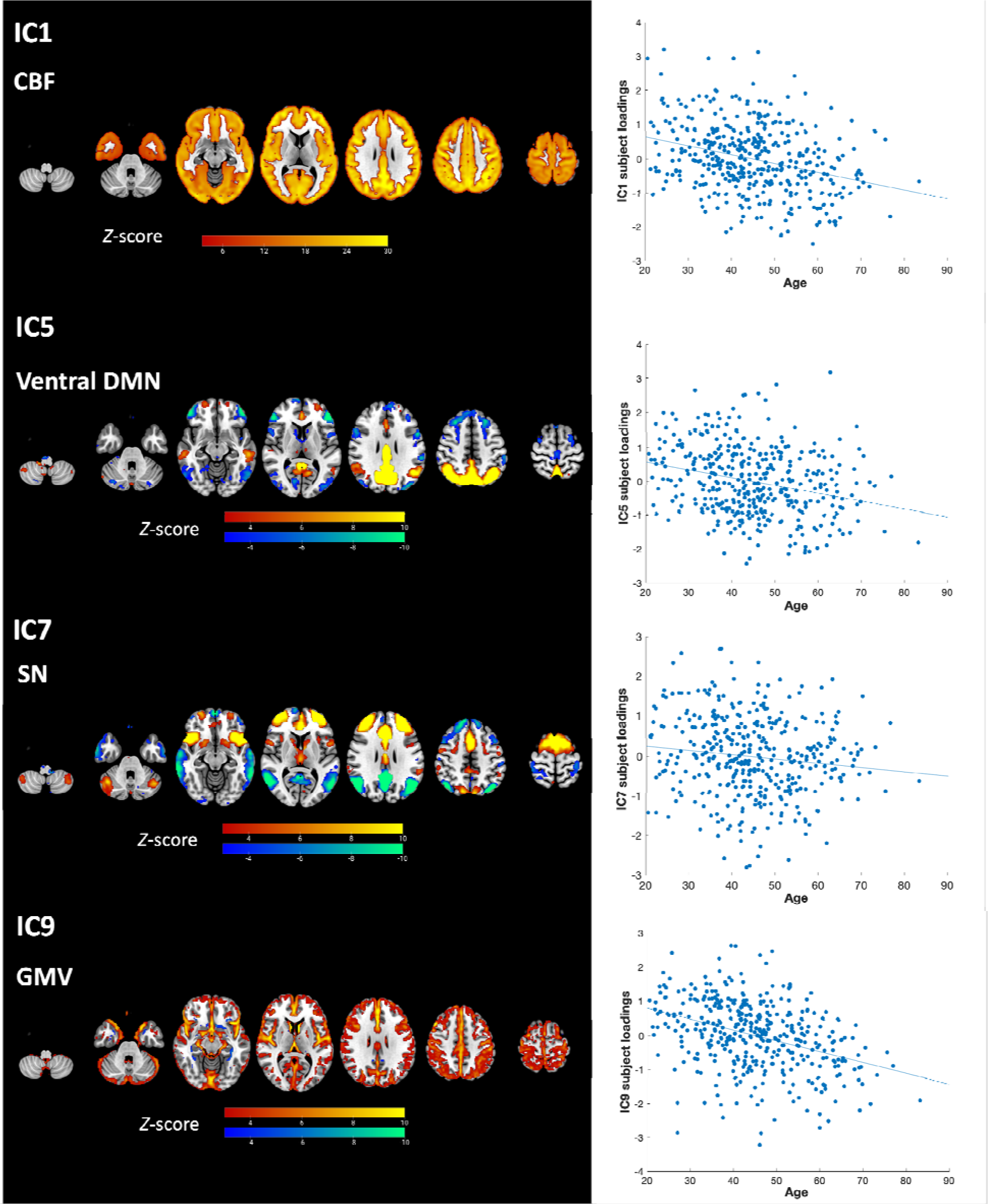
Brain visualization and scatter plots of subject loadings against age of the linked independent component analysis components (ICs) showing strong age effects.

**Figure 5.**
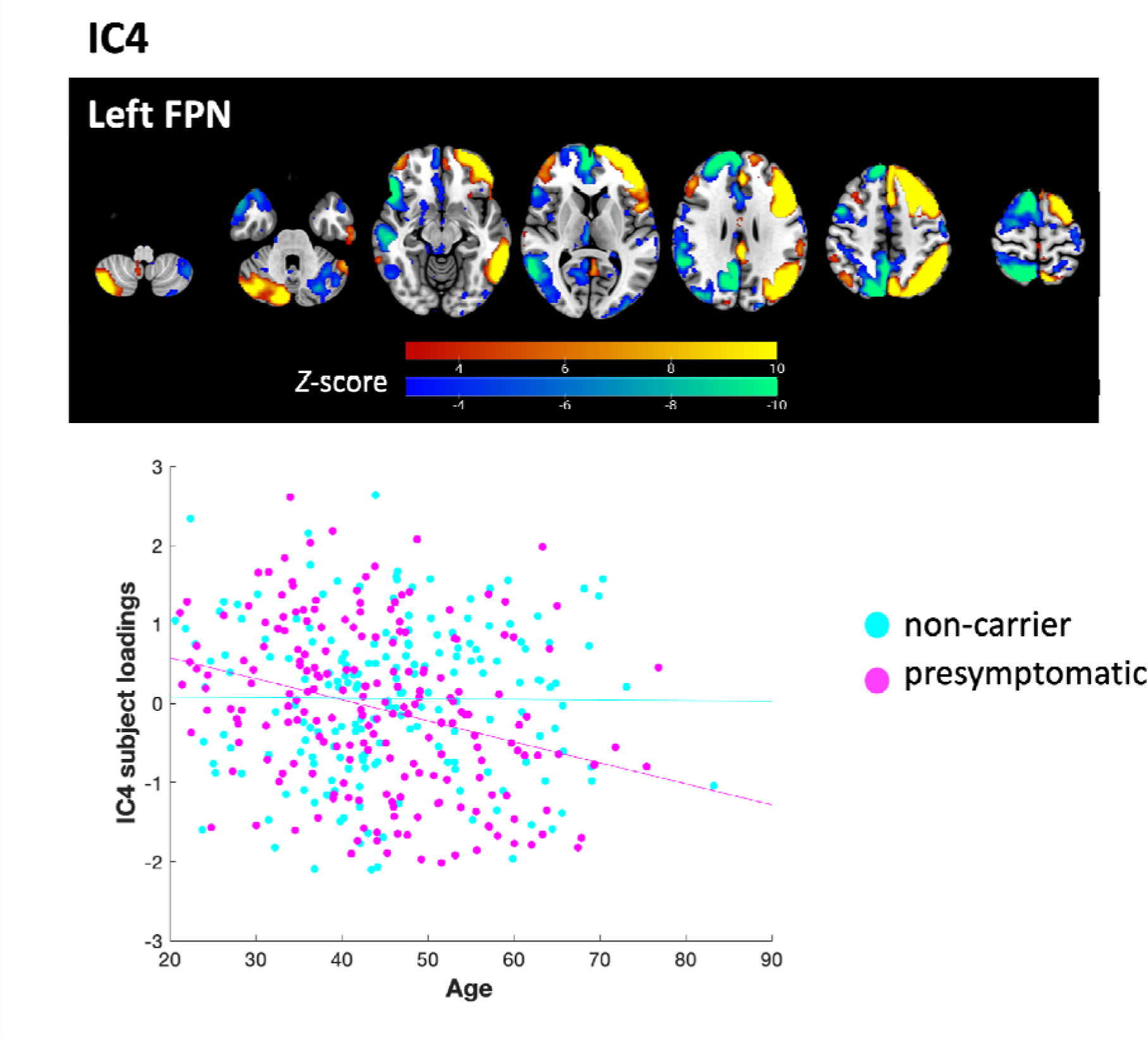
Brain visualization of linked independent component analysis component 4 (IC4), showing the left frontoparietal network (FPN). For visualization, the brain spatial map threshold is set to 3 < |Z| < 10. The scatter plot shows the correlation between age and IC4 subject loading , separated by pre-symptomatic genetic mutation carriers (r = -0.30, *P* < 0.001) and non-carriers (r = -0.0087, *P* = 0.91).

**Table 2.**
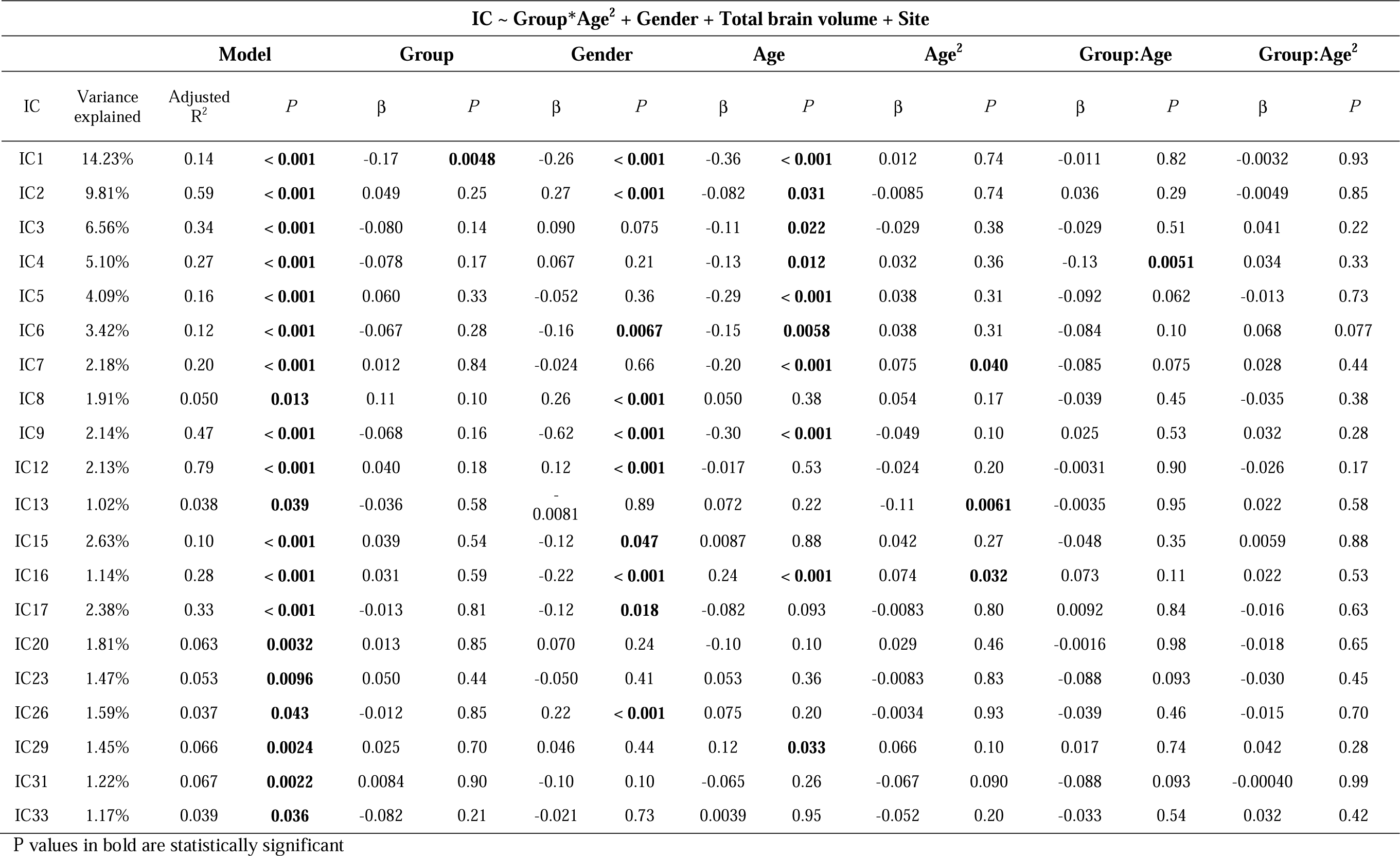
Multiple regression results of the linked independent component analysis components of interest (IC).

### 3.4. Relationship between neuroimaging components and cognitive function

All linked ICA components that showed cognition-related differences between the two groups reflected a single neuroimaging modality. No component showed a different association with cognition component 1 between non-carriers and pre-symptomatic mutation carriers (**Supplementary Table 1**).

In regards to component 2 (**Supplementary Table 2**), IC2, indicating global GMV, showed an interaction with genetic mutation in predicting cognition component 2 (IC:Group t = -2.73, *P* = 0.0066): non-carriers showed a positive association between IC2 subject loadings and good performance on executive functions and poor performance on visuospatial memory tasks (r = 0.17, *P* = 0.026), while this association was not significant in pre-symptomatic mutation carriers (r = -0.12, *P* = 0.10). There was a significant 3-way interaction between group, age and IC subject loadings of the left frontoparietal network (i.e., IC4, IC:Group:Age^2 t = -2.20, *P* = 0.029) in predicting cognition component 2. Visualizing the results (**Figure 6**) indicates that as age increased, an increased association between the left frontoparietal network and good performance on executive functions and poor performance on visuospatial memory tasks was found in pre-symptomatic genetic mutation carriers. This result was confirmed in a post-hoc test showing that a significant two-way interaction between IC4 and age in predicting these cognitive performances was found in pre-symptomatic genetic mutation carriers (IC:Age^2 t = -2.14, *P* = 0.033) but not in non-carriers (IC:Age^2 t = 1.70, *P* = 0.090). Significant 3-way interactions (IC:Group:Age^2) were also observed for the component of ventral default mode network (IC5, t = -2.73, *P* = 0.0068) and salience network (IC7, t = -3.14, *P* = 0.0018). The effects in both components suggested an age-varying association between network activity and performance on executive functions and visuospatial memory in non-carriers but not in pre-symptomatic mutation carriers (**Figure 6**).

**Figure 6.**
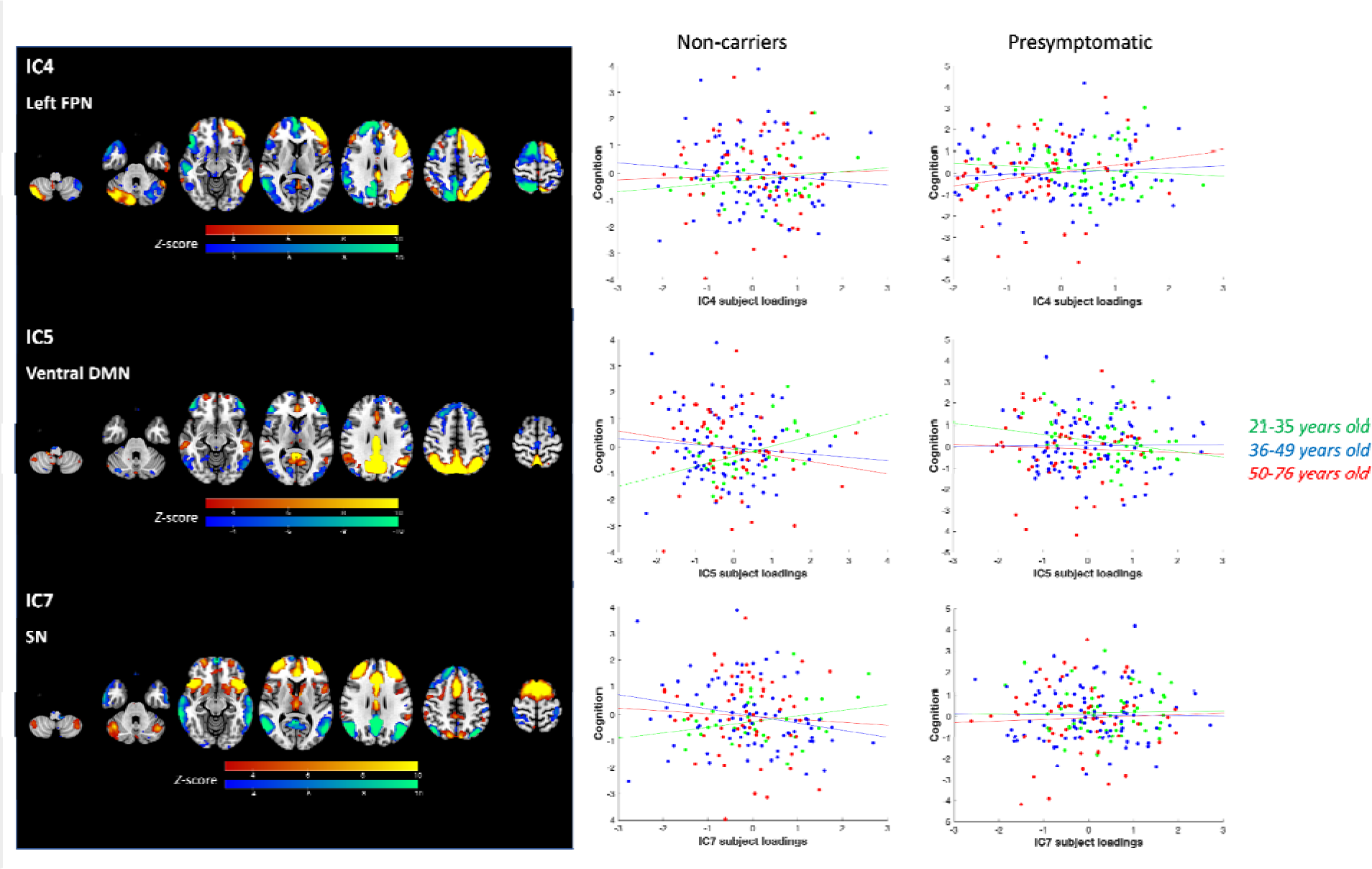
Linked independent component analysis components showing three-way interactions between subject loadings with group (genetic mutation) and age in predicting cognition component 2. IC4 indicates the left frontoparietal network (FPN), IC5 indicates the ventral default mode network (DMN), and IC7 indicates the salience network (SN). The brain visualization and scatter plots are shown. The scatter plots show the correlation between IC subject loading scores and PCA cognition component 2 scores, for visualization purpose separated by pre-symptomatic genetic mutation carriers and non-carriers and three age groups.

In a post-hoc analysis to examine the relationship between age and executive functions, which are the most commonly affected cognitive domains in FTD, we have selected only the tests examining executive functions, attention, and processing speed and performed a PCA on them (**Supplementary** Figure 2). We examined the relationship between age and the significant PCA component (i.e., principal component 1) representing the overall performance of these tests. Results showed a negative association between age and this component in both pre-symptomatic mutation carriers (Age t = -6.78, *P* < 0.001; Age^2 t = -2.73, *P* = 0.007) and non-carriers (Age t = -4.21, *P* < 0.001; Age^2 t = -3.37, *P* < 0.001).

## 4. Discussion

In this study, we co-analyzed GMV, CBF, and functional network integrity. Interplay across modalities did not relate to genetic groups or cognition. Pre-symptomatic genetic mutation carriers showed a decrease with age in the left frontoparietal network integrity while non-carriers did not, suggesting a gene-related neurodegenerative consequence above normal ageing. Executive functions of pre-symptomatic mutation carriers dissociated from the level of atrophy but became dependent on the left frontoparietal network integrity with age. These results suggest that maintaining frontoparietal network integrity may support cognitive function despite age-related atrophy and hypoperfusion in pre-symptomatic FTD.

The age-related decreases in CBF and default mode network activity found in this study are consistent with the commonly observed changes in perfusion [51, 52] and functional network [53] in normal aging. Global GMV also decreased with age, consistent with findings from previous multimodal neuroimaging fusion studies [44, 54] and normal aging pattern of the brain [55]. The component representing global GMV (IC2) in this study did not significantly differ between pre-symptomatic mutation carriers and non-carriers accounting for age effects. The main reason may be this component is dominated by the effect of ageing, as linked ICA identifies components in a data-driven manner. Signals in this component are mostly influenced by age-related variances, which can be attributed to the wide age range of participants, spanning from 20 to 83 years old. On the other hand, studies employing hypothesis-driven approaches identified atrophy patterns that are optimised to detect pre-symptomatic differences [4, 7]. Thus, the difference in atrophy patterns identified in those studies might be specific to pre-symptomatic mutation carriers versus age-matched controls [7, 56], while IC2 in our study predominantly reflects age-related atrophy as reported in previous studies [44, 54, 57].

More importantly, we illustrated the age- and cognition-relevant divergence of frontoparietal network integrity between pre-symptomatic mutation carriers and non-carriers. Pre-symptomatic mutation carriers showed a decrease in the left frontoparietal network integrity with age while non-carriers did not, suggesting that the lateralized frontoparietal network is the functional network most affected by FTD mutations with age. Salience network connectivity is commonly reduced in symptomatic behavioural variant FTD (bvFTD) and associated with disease severity [46, 58], but remains unchanged at the pre-symptomatic stage [59]. Altered default mode network connectivity has been found in both pre-symptomatic *MAPT* mutation carriers and bvFTD subjects [46, 59]. In this study, we did not find the default mode network or the salience network significantly different between genetic mutation carriers and non-carriers when accounting for the age effects. Nevertheless, when relating to executive function, attention and processing speed, the associations between the ventral default mode network and the salience network, respectively, with performance on these functions were found particularly in younger non-carriers but not in pre-symptomatic mutation carriers, suggesting the cognitive reliance on these functional networks breaks down in genetic mutation carriers and during ageing. Understanding such an effect would be important for gaining insights into the mechanisms of cognitive decline and the maintenance of executive functions especially at the pre-symptomatic stage of cognitive impairment.

Pre-symptomatic mutation carriers maintain similar global cognition to non-carriers. However, we demonstrated that global cognition showed a trend of more rapid decline with age in pre-symptomatic mutation carriers from the age of early 20s to 80s. We found no significant association between age and executive function, attention, and processing speed in either groups, contrasting previous reports of age-related declines in executive functions [60], potentially due to different analysis methods. Here, executive functions were represented by the second principal component, which should be interpreted in relation to the negatively loading visuospatial memory and in the context of the first principal component. Principal component 1 captured the well-documented age-related decline in global cognitive functions including executive functions and memory [61, 62]. Principal component 2 may represent aspects of executive functions, that are independent or orthogonal to the general cognitive decline, possibly reflecting individual variability specific to the cognitive tests. Hence, the age-related differences of these specific executive functions might be moderated by the age-related effect of visuospatial memory, while executive functions declining alongside memory are likely already captured by principal component 1. Post-hoc analysis showed a negative age-executive functions association, consistent with reported age-related executive declines [60]. The performance related to attention, processing speed and executive function correlated with global GMV in non-carriers, while correlated with left frontoparietal network integrity in pre-symptomatic mutation carriers especially as they get older. It suggests that in genetic mutation carriers, executive functions dissociated from GMV and were maintained by frontoparietal network integrity. Frontoparietal network is important for cognitive flexibility especially for executive function [63, 64], which is one of the most commonly affected cognitive domains in FTD [1]. A recent study has found that pre-symptomatic *C9orf72* mutation carriers showed lower attention and executive function compared to non-carriers [62]. Our study provides further evidence suggesting that these cognitive domains are sensitive to alternations at the earlier stage of the disease. Given that CBF and GMV significantly decreased with age regardless of genetic mutations, and the reliance on other functional networks for cognitive performance broke down in genetic mutation carriers, maintaining frontoparietal network integrity might be the key to slowing cognitive decline at the pre-symptomatic stage of FTD, particularly slowing the decline in executive functions.

The atrophy patterns can be different across different genetic mutations. The *GRN* genetic mutation is known for causing asymmetric atrophy while the atrophy patterns of FTD associated with *MAPT* genetic mutation are typically symmetric [7, 56, 65]. We observed asymmetric relationship between functional network integrity and age in the *GRN* mutation carriers, indicating that the asymmetric vulnerability to genetic mutation can be manifested at the pre-symptomatic stage. Specifically, we observed a relationship between age and the left frontoparietal network in the *GRN* mutation carriers, although the lack of significance in other genetic groups may be attributed to smaller sample sizes compared to *GRN* mutation carriers. Such finding is consistent with previous studies showing selective vulnerability of the left hemisphere [56, 66, 67]. Moreover, there is inherent asymmetry in several human cognitive systems, including language and executive functions which could be significantly impaired in FTD [68-70]. Although the cellular mechanisms of selective vulnerability are not well understood, it would be important to investigate the laterality of changes in future studies, especially considering the dynamical interactions between brain networks which shape cognition.

This study benefits from pathological confidence arising from genetic characterisation, and the large sample size of pre-symptomatic mutation carriers through the multi-center GENFI study. This study combines GMV, CBF and functional networks in pre-symptomatic FTD genetic mutation carriers. Linking neurobiological changes is important given potential synergistic effects. Although, we found no interplay across modalities, relating frontoparietal network to other unexplored pathologies like tau, amyloid and neurotransmitters may prove informative [46, 59, 71, 72], given its age- and cognition-related distinctions between genetic mutation carriers and non-carriers observed in our study.

The study also has limitations. First, the variability of MRI acquisition scanners and sequences through the multi-center cohort is higher than a single-center study. However, we mitigated the effects through the use of the normalization, denoising and statistical adjustment for site effects. We recognize that multi-center and multi-scanner correction for ASL could potentially be improved. A standard approach would be the use of flow phantoms for calibrating a scanner’s ASL signal to a ground-truth flow rate [73]. Currently, however, this is not implemented in most ASL studies. Existing methods of pre-model (e.g., using ExploreASL) or within-model corrections [74] along with data-driven and model-driven corrections for sites and scanners remain the most pragmatic approach. Second, this study is cross-sectional. This should be noted when interpreting age effects, as dynamic ageing effects require longitudinal data. More follow-up visits of the ongoing GENFI cohort will allow a longitudinal examination of these participants. Third, only adults were included thus potentially missing the changes manifested before adulthood caused by genetic mutation. A new cohort within GENFI is starting which aims to study family members below the age of 18. Finally, our current study focused on integrating spatial maps of network activity in relation to atrophy and perfusion. However, functional connectivity between networks is another important factor to be considered [4]. The joint consideration of activity and connectivity might better characterize brain dynamics and cognitive performance [75]. Therefore, future research could investigate the intercorrelations between functional connectivity and multiple neuroimaging modalities or integrate time-course functional data with spatial maps from other modalities.

In conclusion, we demonstrated that the frontoparietal network integrity might support cognitive function despite atrophy and hypoperfusion in pre-symptomatic FTD. While the linked ICA components indicating an interplay across modalities did not relate to genetic groups and cognition, linking neuroimaging, especially functional network integrity, with other neuropathological changes may be a future direction for genetic FTD at the pre-symptomatic stage. The dissociation of changes in structure, perfusion and network activity in pre-symptomatic FTD has implications for clinical trials design, and strategies for prevention or treatments for nominally well people at high risk of FTD.

## Data Availability

All data produced in the present study are available upon reasonable request to the authors. Data restrictions may apply in order to preserve participant confidentiality.

## Consent Statement

All participants provided informed consent.

## Acknowledgements

X.L. is supported by the Cambridge Commonwealth, European and International Trust. J.C.V.S., L.C.J. and H.S. are supported by the Dioraphte Foundation grant 09-02-03-00, Association for Frontotemporal Dementias Research Grant 2009, Netherlands Organization for Scientific Research grant HCMI 056-13-018, ZonMw Memorabel (Deltaplan Dementie, project number 733 051 042), ZonMw Onderzoeksprogramma Dementie (YOD-INCLUDED, project number10510032120002), EU Joint Programme-Neurodegenerative Disease Research-GENFI-PROX, Alzheimer Nederland and the Bluefield Project. R.S-V. is supported by Alzheimer’s Research UK Clinical Research Training Fellowship (ARUK-CRF2017B-2) and has received funding from Fundació Marató de TV3, Spain (grant no. 20143810). C.G. received funding from EU Joint Programme-Neurodegenerative Disease Research-Prefrontals Vetenskapsrådet Dnr 529-2014-7504, EU Joint Programme-Neurodegenerative Disease Research-GENFI-PROX, Vetenskapsrådet 2019-0224, Vetenskapsrådet 2015-02926, Vetenskapsrådet 2018-02754, the Swedish FTD Inititative-Schörling Foundation, Alzheimer Foundation, Brain Foundation, Dementia Foundation and Region Stockholm ALF-project. D.G. received support from the EU Joint Programme-Neurodegenerative Disease Research and the Italian Ministry of Health (PreFrontALS) grant 733051042. R.V. has received funding from the Mady Browaeys Fund for Research into Frontotemporal Dementia. J.L. received funding for this work by the Deutsche Forschungsgemeinschaft German Research Foundation under Germany’s Excellence Strategy within the framework of the Munich Cluster for Systems Neurology (EXC 2145 SyNergy—ID 390857198). M.O. has received funding from Germany’s Federal Ministry of Education and Research (BMBF). E.F. has received funding from a Canadian Institute of Health Research grant #327387. M.M. has received funding from a Canadian Institute of Health Research operating grant and the Weston Brain Institute and Ontario Brain Institute. F.M. is supported by the Tau Consortium and has received funding from the Carlos III Health Institute (PI19/01637). J.D.R. is supported by the Bluefield Project and the National Institute for Health and Care Research University College London Hospitals Biomedical Research Centre, and has received funding from an MRC Clinician Scientist Fellowship (MR/M008525/1) and a Miriam Marks Brain Research UK Senior Fellowship. Several authors of this publication (J.C.V.S., M.S., R.V., A.d.M., M.O., R.V., J.D.R.) are members of the European Reference Network for Rare Neurological Diseases (ERN-RND) - Project ID No 739510. K.A.T. was supported by the Guarantors of Brain (G101149) and Alzheimer’s Society (Grant Nr. 602). J.B.R. has received funding from the Welcome Trust (103838; 220258) and is supported by the Cambridge University Centre for Frontotemporal Dementia, the Medical Research Council (MC_UU_00030/14; MR/T033371/1) and the National Institute for Health Research Cambridge Biomedical Research Centre (NIHR203312: BRC-1215-20014) and the Holt Fellowship. This work was also supported by the EU Joint Programme-Neurodegenerative Disease Research GENFI-PROX grant [2019-02248; to J.D.R., M.O., B.B., C.G., J.C.V.S. and M.S. For the purpose of open access, the author has applied a CC BY public copyright licence to any Author Accepted Manuscript version arising from this submission.

## The GENFI Consortium Author List

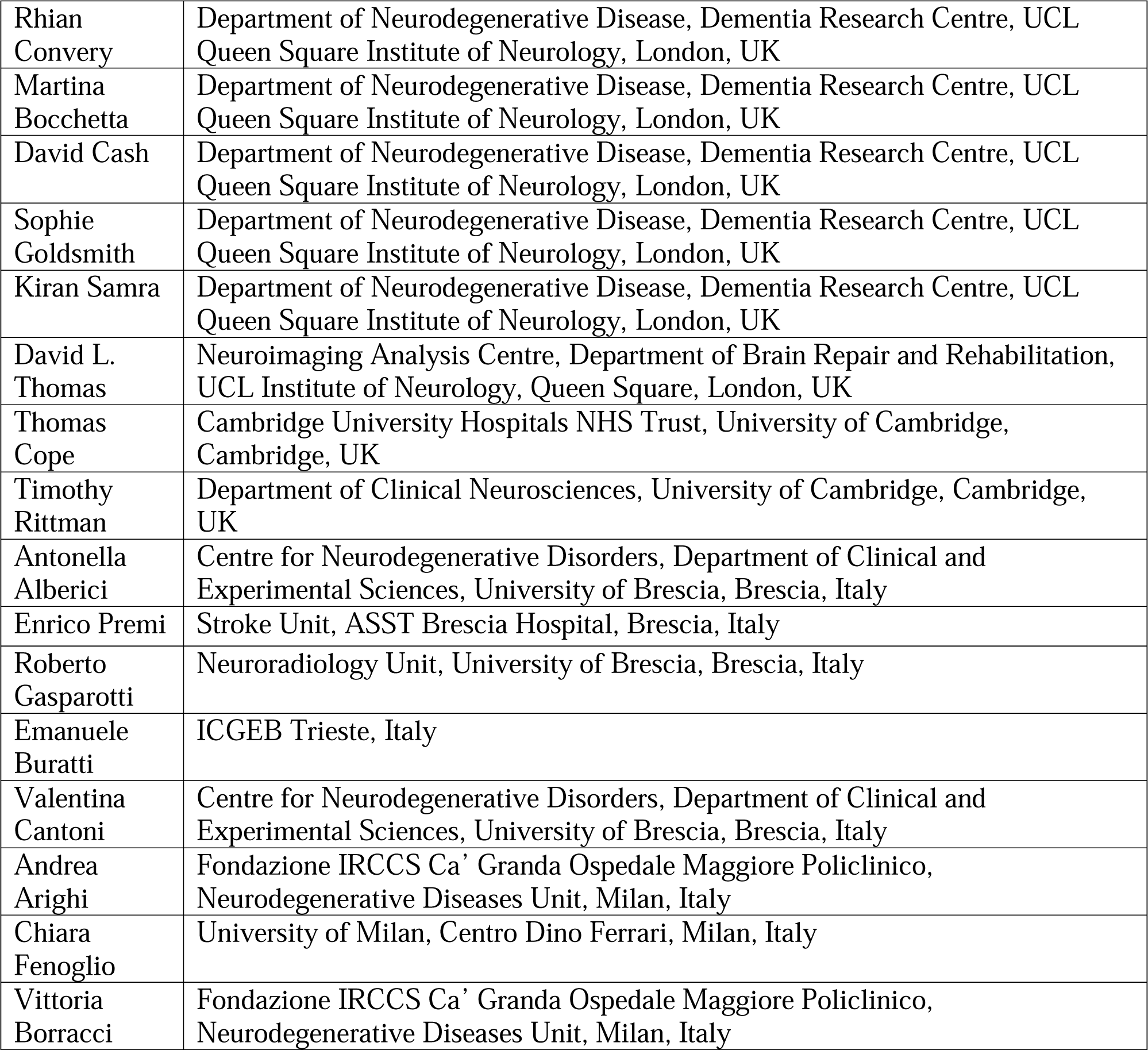
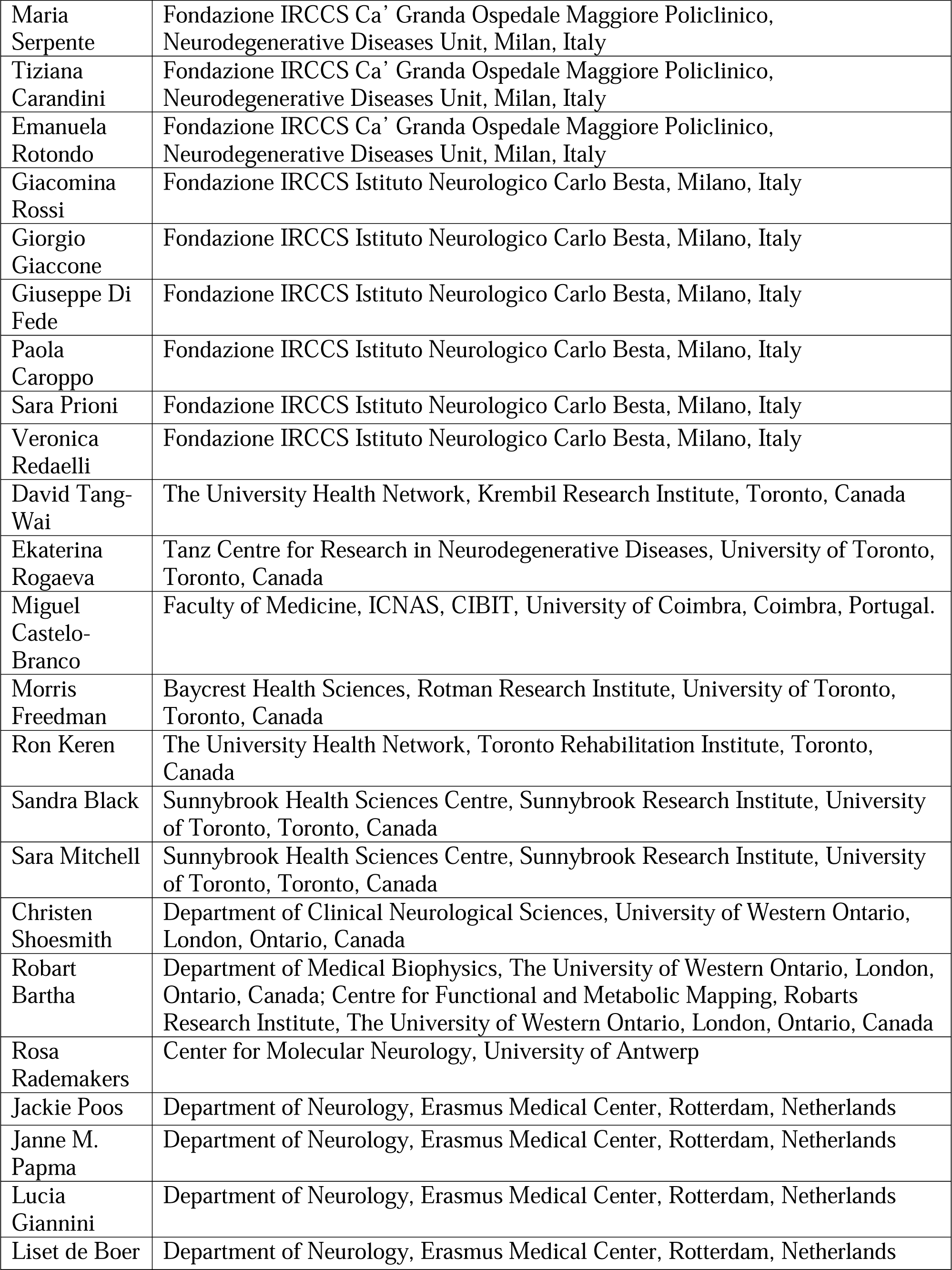
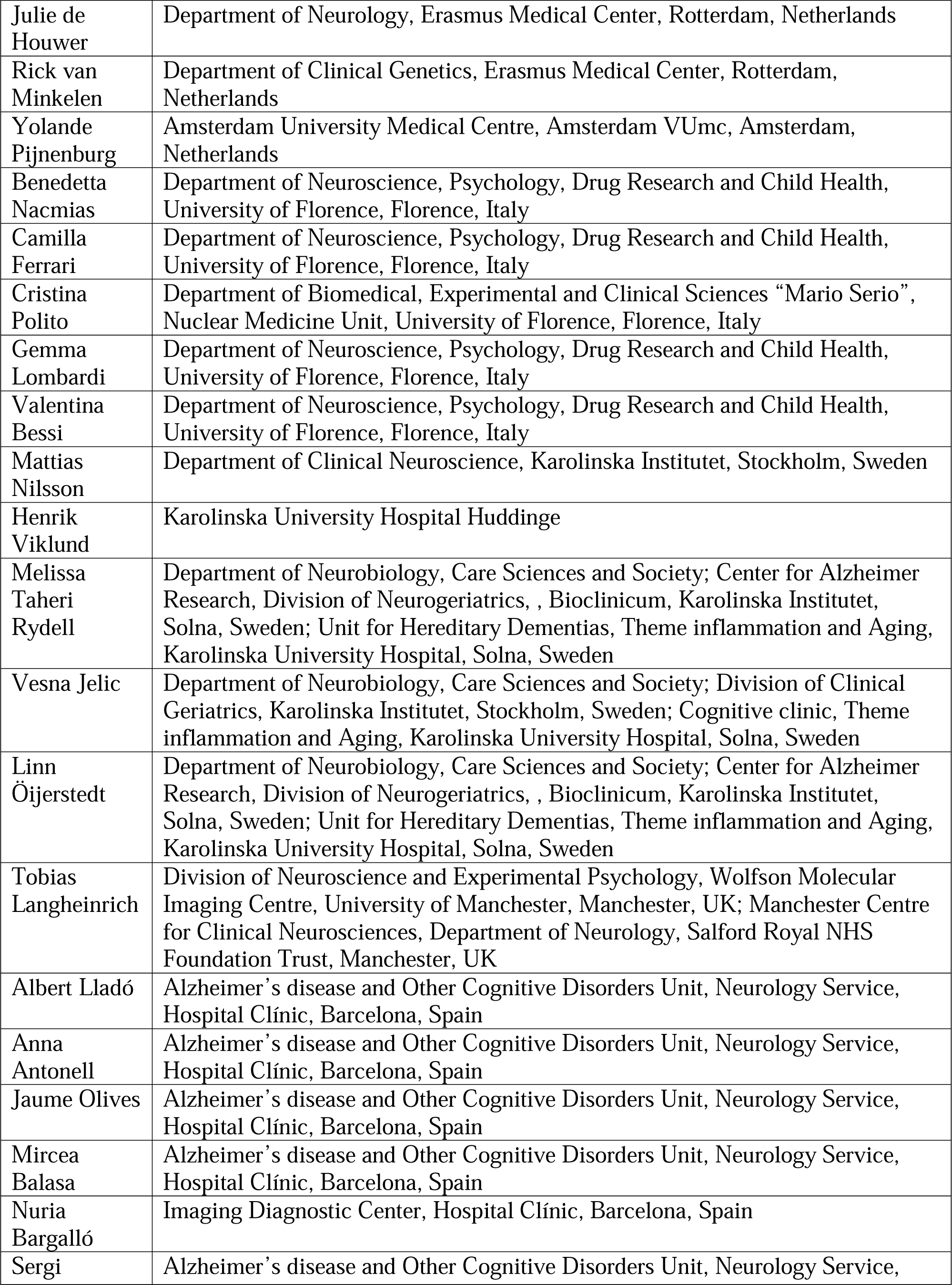
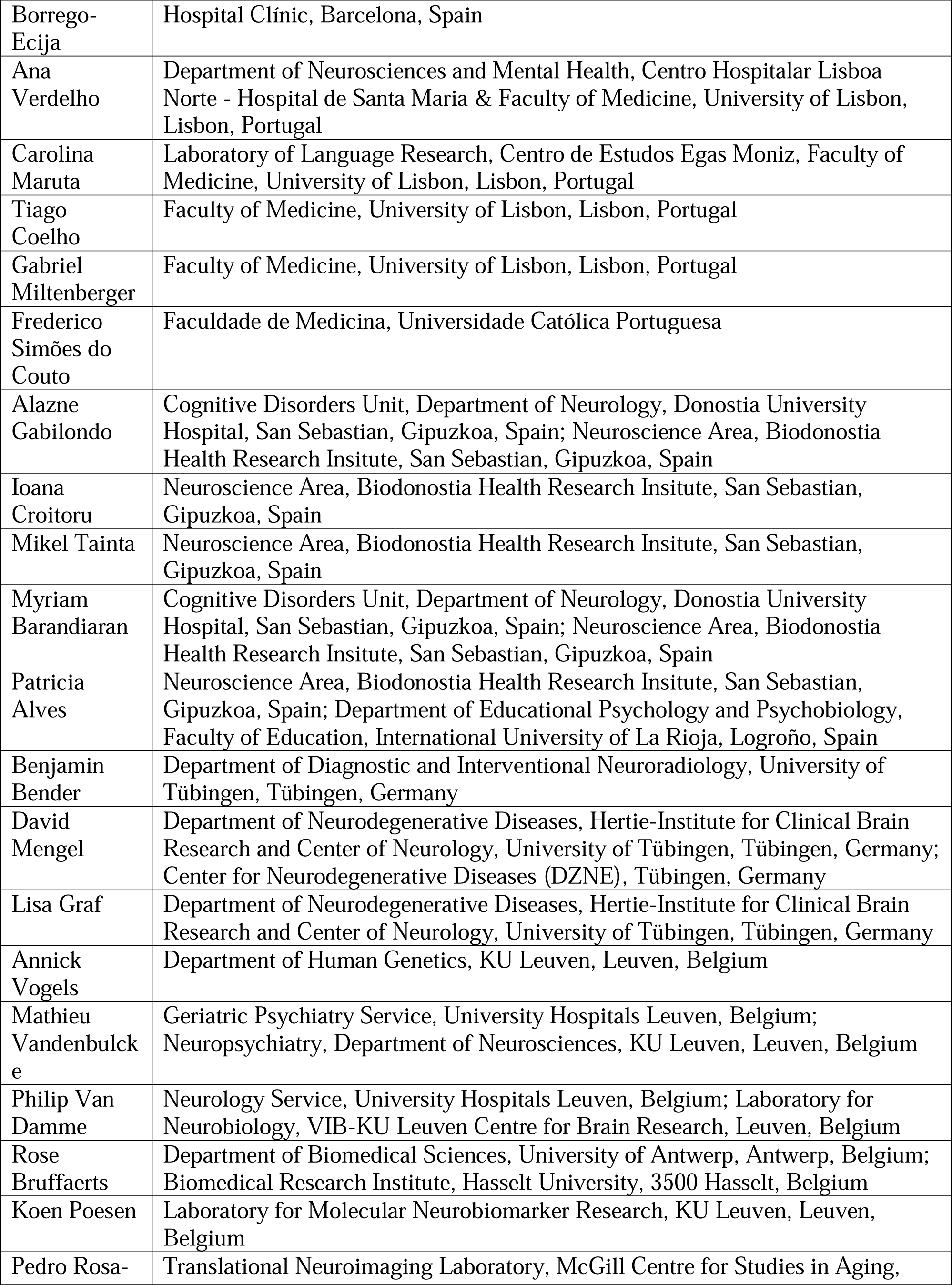
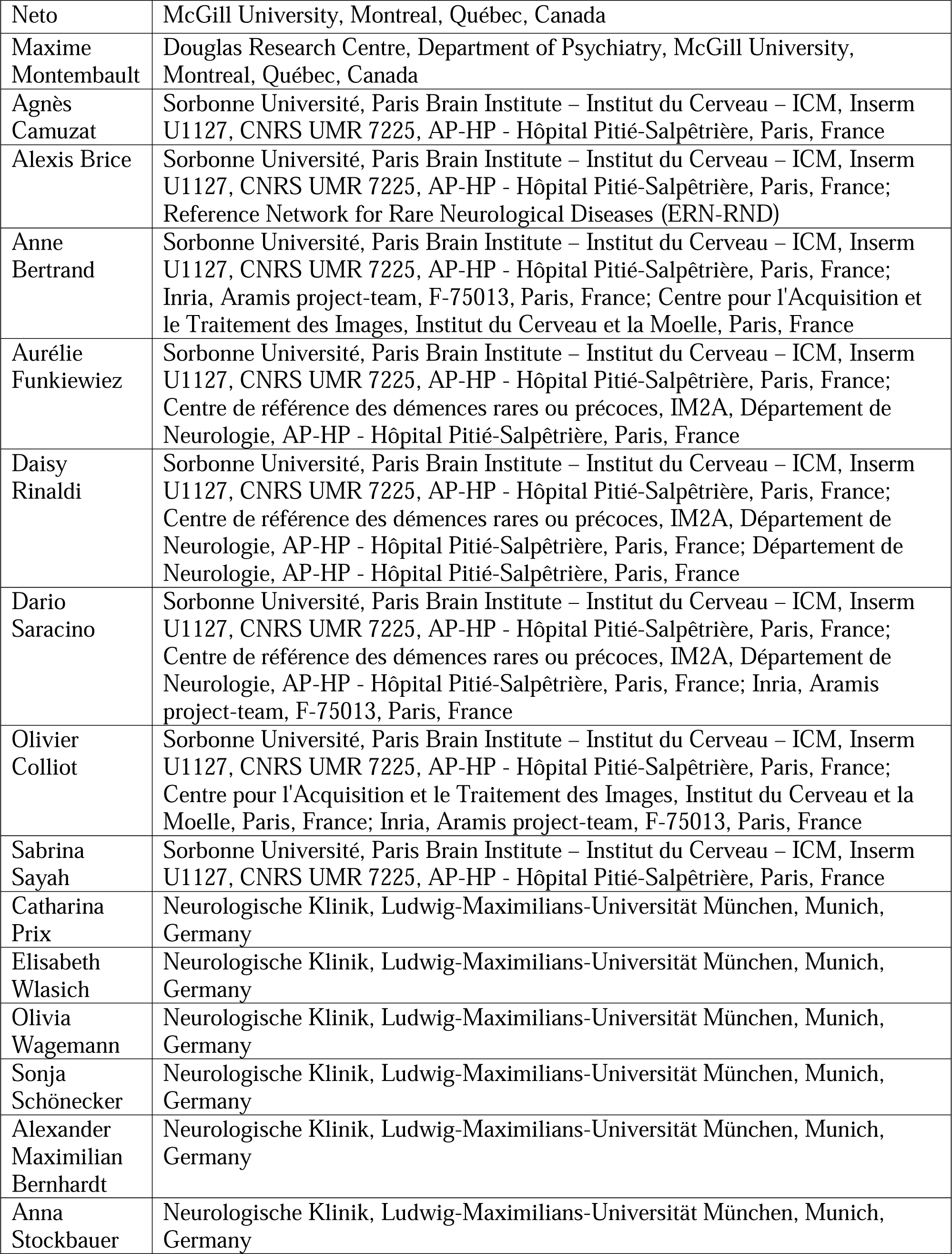
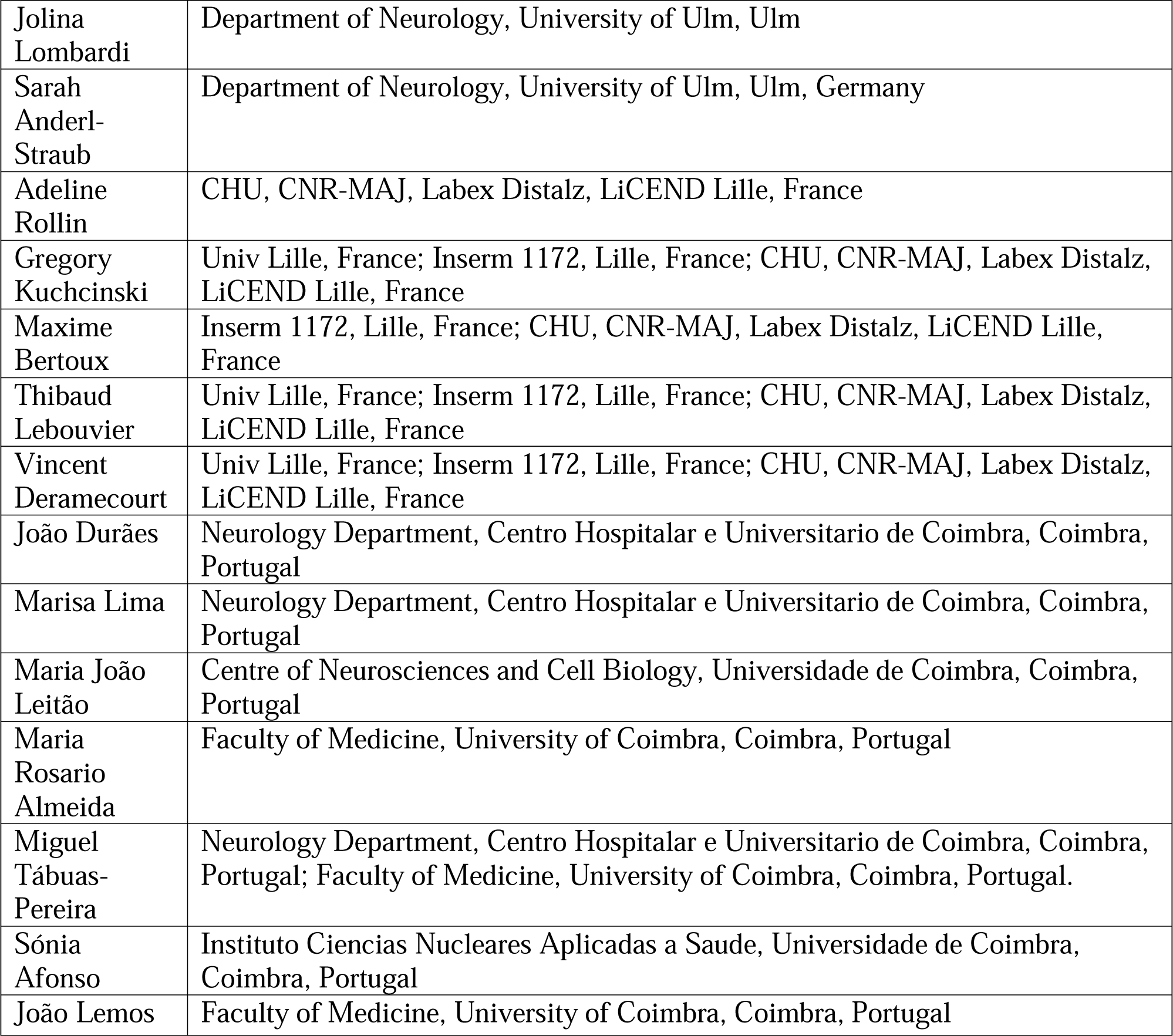

## References

[1] Bang J, Spina S, Miller BL. Frontotemporal dementia. Lancet. 2015;386:1672–82.

[2] Rohrer JD, Guerreiro R, Vandrovcova J, Uphill J, Reiman D, Beck J, et al. The heritability and genetics of frontotemporal lobar degeneration. Neurology. 2009;73:1451–6.

[3] Kinnunen KM, Cash DM, Poole T, Frost C, Benzinger TLS, Ahsan RL, et al. Presymptomatic atrophy in autosomal dominant Alzheimer’s disease: A serial magnetic resonance imaging study. Alzheimers Dement. 2018;14:43–53.

[4] Tsvetanov KA, Gazzina S, Jones PS, van Swieten J, Borroni B, Sanchez-Valle R, et al. Brain functional network integrity sustains cognitive function despite atrophy in presymptomatic genetic frontotemporal dementia. Alzheimers Dement. 2021;17:500–14.

[5] Jack CR, Jr., Knopman DS, Jagust WJ, Shaw LM, Aisen PS, Weiner MW, et al. Hypothetical model of dynamic biomarkers of the Alzheimer’s pathological cascade. Lancet Neurol. 2010;9:119–28.

[6] Rohrer JD, Nicholas JM, Cash DM, van Swieten J, Dopper E, Jiskoot L, et al. Presymptomatic cognitive and neuroanatomical changes in genetic frontotemporal dementia in the Genetic Frontotemporal dementia Initiative (GENFI) study: a cross-sectional analysis. Lancet Neurol. 2015;14:253–62.

[7] Cash DM, Bocchetta M, Thomas DL, Dick KM, van Swieten JC, Borroni B, et al. Patterns of gray matter atrophy in genetic frontotemporal dementia: results from the GENFI study. Neurobiology of Aging. 2018;62:191–6.

[8] Mutsaerts H, Mirza SS, Petr J, Thomas DL, Cash DM, Bocchetta M, et al. Cerebral perfusion changes in presymptomatic genetic frontotemporal dementia: a GENFI study. Brain. 2019;142:1108–20.

[9] Rittman T, Borchert R, Jones S, van Swieten J, Borroni B, Galimberti D, et al. Functional network resilience to pathology in presymptomatic genetic frontotemporal dementia. Neurobiol Aging. 2019;77:169–77.

[10] Whiteside DJ, Malpetti M, Jones PS, Ghosh BCP, Coyle-Gilchrist I, van Swieten JC, et al. Temporal dynamics predict symptom onset and cognitive decline in familial frontotemporal dementia. Alzheimers Dement. 2022.

[11] Miyagawa T, Brushaber D, Syrjanen J, Kremers W, Fields J, Forsberg LK, et al. Utility of the global CDR((R)) plus NACC FTLD rating and development of scoring rules: Data from the ARTFL/LEFFTDS Consortium. Alzheimers Dement. 2020;16:106–17.

[12] Gorno-Tempini ML, Hillis AE, Weintraub S, Kertesz A, Mendez M, Cappa SF, et al. Classification of primary progressive aphasia and its variants. Neurology. 2011;76:1006–14.

[13] Rascovsky K, Hodges JR, Knopman D, Mendez MF, Kramer JH, Neuhaus J, et al. Sensitivity of revised diagnostic criteria for the behavioural variant of frontotemporal dementia. Brain. 2011;134:2456–77.

[14] Mioshi E, Hsieh S, Savage S, Hornberger M, Hodges JR. Clinical staging and disease progression in frontotemporal dementia. Neurology. 2010;74:1591–7.

[15] Wear HJ, Wedderburn CJ, Mioshi E, Williams-Gray CH, Mason SL, Barker RA, Hodges JR. The Cambridge Behavioural Inventory revised. Dement Neuropsychol. 2008;2:102–7.

[16] Morris JC, Weintraub S, Chui HC, Cummings J, Decarli C, Ferris S, et al. The Uniform Data Set (UDS): clinical and cognitive variables and descriptive data from Alzheimer Disease Centers. Alzheimer Dis Assoc Disord. 2006;20:210–6.

[17] Corrigan JD, Hinkeldey NS. Relationships between parts A and B of the Trail Making Test. J Clin Psychol. 1987;43:402–9.

[18] Delis DC KE, Kramer J, den Buysch HO, Noens ILJ, Berckelaer-Onnes IA. DKEFS: Delis-Kaplan Executive Function System: Color-Word Interference Test: Handleiding: Pearson; 2008.

[19] Moore K, Convery R, Bocchetta M, Neason M, Cash DM, Greaves C, et al. A modified Camel and Cactus Test detects presymptomatic semantic impairment in genetic frontotemporal dementia within the GENFI cohort. Appl Neuropsychol Adult. 2022;29:112–9.

[20] Tombaugh TN, Kozak J, Rees L. Normative data stratified by age and education for two measures of verbal fluency: FAS and animal naming. Arch Clin Neuropsychol. 1999;14:167–77.

[21] Poos JM, Russell LL, Peakman G, Bocchetta M, Greaves CV, Jiskoot LC, et al. Impairment of episodic memory in genetic frontotemporal dementia: A GENFI study. Alzheimers Dement (Amst). 2021;13:e12185.

[22] Ashburner J. A fast diffeomorphic image registration algorithm. Neuroimage. 2007;38:95–113.

[23] Mutsaerts H, Petr J, Thomas DL, De Vita E, Cash DM, van Osch MJP, et al. Comparison of arterial spin labeling registration strategies in the multi-center GENetic frontotemporal dementia initiative (GENFI). J Magn Reson Imaging. 2018;47:131–40.

[24] Mutsaerts H, Petr J, Groot P, Vandemaele P, Ingala S, Robertson AD, et al. ExploreASL: An image processing pipeline for multi-center ASL perfusion MRI studies. Neuroimage. 2020;219:117031.

[25] Mutsaerts HJ, Petr J, Vaclavu L, van Dalen JW, Robertson AD, Caan MW, et al. The spatial coefficient of variation in arterial spin labeling cerebral blood flow images. J Cereb Blood Flow Metab. 2017;37:3184–92.

[26] Asllani I, Borogovac A, Brown TR. Regression algorithm correcting for partial volume effects in arterial spin labeling MRI. Magnetic Resonance in Medicine. 2008;60:1362–71.

[27] Pasternak M, Mirza SS, Luciw N, Mutsaerts HJMM, Petr J, Thomas D, et al. Longitudinal cerebral perfusion in presymptomatic genetic frontotemporal dementia: GENFI results. Alzheimer’s & Dementia. 2024;20:3525–42.

[28] Li H, Smith SM, Gruber S, Lukas SE, Silveri MM, Hill KP, et al. Denoising scanner effects from multimodal MRI data using linked independent component analysis. Neuroimage. 2020;208:116388.

[29] Chen J, Liu J, Calhoun VD, Arias-Vasquez A, Zwiers MP, Gupta CN, et al. Exploration of scanning effects in multi-site structural MRI studies. J Neurosci Methods. 2014;230:37–50.

[30] Ashburner J, Friston KJ. Diffeomorphic registration using geodesic shooting and Gauss– Newton optimisation. NeuroImage. 2011;55:954–67.

[31] Shirzadi Z, Crane DE, Robertson AD, Maralani PJ, Aviv RI, Chappell MA, et al. Automated removal of spurious intermediate cerebral blood flow volumes improves image quality among older patients: A clinical arterial spin labeling investigation. Journal of Magnetic Resonance Imaging. 2015;42:1377–85.

[32] Alsop DC, Detre JA, Golay X, Günther M, Hendrikse J, Hernandez-Garcia L, et al. Recommended implementation of arterial spin-labeled perfusion MRI for clinical applications: A consensus of the ISMRM perfusion study group and the European consortium for ASL in dementia. Magnetic Resonance in Medicine. 2015;73:102–16.

[33] Cusack R, Vicente-Grabovetsky A, Mitchell DJ, Wild CJ, Auer T, Linke AC, Peelle JE. Automatic analysis (aa): efficient neuroimaging workflows and parallel processing using Matlab and XML. Front Neuroinform. 2014;8:90.

[34] Pruim RHR, Mennes M, van Rooij D, Llera A, Buitelaar JK, Beckmann CF. ICA-AROMA: A robust ICA-based strategy for removing motion artifacts from fMRI data. Neuroimage. 2015;112:267–77.

[35] Geerligs L, Tsvetanov KA, Cam C, Henson RN. Challenges in measuring individual differences in functional connectivity using fMRI: The case of healthy aging. Hum Brain Mapp. 2017;38:4125–56.

[36] Calhoun VD, Adali T, Pearlson GD, Pekar JJ. A method for making group inferences from functional MRI data using independent component analysis. Hum Brain Mapp. 2001;14:140–51.

[37] McKeown MJ, Makeig S, Brown GG, Jung TP, Kindermann SS, Bell AJ, Sejnowski TJ. Analysis of fMRI data by blind separation into independent spatial components. Hum Brain Mapp. 1998;6:160–88.

[38] Rosazza C, Minati L. Resting-state brain networks: literature review and clinical applications. Neurol Sci. 2011;32:773–85.

[39] Beckmann CF, DeLuca M, Devlin JT, Smith SM. Investigations into resting-state connectivity using independent component analysis. Philos Trans R Soc Lond B Biol Sci. 2005;360:1001–13.

[40] Damoiseaux JS, Rombouts SA, Barkhof F, Scheltens P, Stam CJ, Smith SM, Beckmann CF. Consistent resting-state networks across healthy subjects. Proc Natl Acad Sci U S A. 2006;103:13848–53.

[41] Smith SM, Fox PT, Miller KL, Glahn DC, Fox PM, Mackay CE, et al. Correspondence of the brain’s functional architecture during activation and rest. Proc Natl Acad Sci U S A. 2009;106:13040–5.

[42] Himberg J, Hyvarinen A. Icasso: software for investigating the reliability of ICA estimates by clustering and visualization. 2003 IEEE XIII Workshop on Neural Networks for Signal Processing (IEEE Cat No03TH8718). 2003:259-68.

[43] Shirer WR, Ryali S, Rykhlevskaia E, Menon V, Greicius MD. Decoding subject-driven cognitive states with whole-brain connectivity patterns. Cereb Cortex. 2012;22:158–65.

[44] Liu X, Tyler LK, Cam CAN, Rowe JB, Tsvetanov KA. Multimodal fusion analysis of functional, cerebrovascular and structural neuroimaging in healthy aging subjects. Hum Brain Mapp. 2022;43:5490–508.

[45] Passamonti L, Tsvetanov KA, Jones PS, Bevan-Jones WR, Arnold R, Borchert RJ, et al. Neuroinflammation and Functional Connectivity in Alzheimer’s Disease: Interactive Influences on Cognitive Performance. The Journal of Neuroscience. 2019;39:7218–26.

[46] Pievani M, de Haan W, Wu T, Seeley WW, Frisoni GB. Functional network disruption in the degenerative dementias. Lancet Neurol. 2011;10:829–43.

[47] Snyder W, Uddin LQ, Nomi JS. Dynamic functional connectivity profile of the salience network across the life span. Hum Brain Mapp. 2021;42:4740–9.

[48] Groves AR, Beckmann CF, Smith SM, Woolrich MW. Linked independent component analysis for multimodal data fusion. Neuroimage. 2011;54:2198–217.

[49] Smith SM, Jenkinson M, Woolrich MW, Beckmann CF, Behrens TE, Johansen-Berg H, et al. Advances in functional and structural MR image analysis and implementation as FSL. Neuroimage. 2004;23 Suppl 1:S208–19.

[50] Wilkinson GN, Rogers CE. Symbolic Description of Factorial Models for Analysis of Variance. Journal of the Royal Statistical Society Series C (Applied Statistics). 1973;22:392–9.

[51] Zhang N, Gordon ML, Goldberg TE. Cerebral blood flow measured by arterial spin labeling MRI at resting state in normal aging and Alzheimer’s disease. Neurosci Biobehav Rev. 2017;72:168–75.

[52] Mokhber N, Shariatzadeh A, Avan A, Saber H, Babaei GS, Chaimowitz G, Azarpazhooh MR. Cerebral blood flow changes during aging process and in cognitive disorders: A review. Neuroradiol J. 2021;34:300–7.

[53] Damoiseaux JS, Beckmann CF, Arigita EJ, Barkhof F, Scheltens P, Stam CJ, et al. Reduced resting-state brain activity in the “default network” in normal aging. Cereb Cortex. 2008;18:1856–64.

[54] Douaud G, Groves AR, Tamnes CK, Westlye LT, Duff EP, Engvig A, et al. A common brain network links development, aging, and vulnerability to disease. Proc Natl Acad Sci U S A. 2014;111:17648–53.

[55] Kennedy KM, Raz N. Normal Aging of the Brain. In: Toga AW, editor. Brain Mapping. Waltham: Academic Press; 2015. p. 603-17.

[56] Fumagalli GG, Basilico P, Arighi A, Bocchetta M, Dick KM, Cash DM, et al. Distinct patterns of brain atrophy in Genetic Frontotemporal Dementia Initiative (GENFI) cohort revealed by visual rating scales. Alzheimers Res Ther. 2018;10:46.

[57] Peelle JE, Cusack R, Henson RN. Adjusting for global effects in voxel-based morphometry: gray matter decline in normal aging. Neuroimage. 2012;60:1503–16.

[58] Zhou J, Greicius MD, Gennatas ED, Growdon ME, Jang JY, Rabinovici GD, et al. Divergent network connectivity changes in behavioural variant frontotemporal dementia and Alzheimer’s disease. Brain. 2010;133:1352–67.

[59] Whitwell JL, Josephs KA, Avula R, Tosakulwong N, Weigand SD, Senjem ML, et al. Altered functional connectivity in asymptomatic MAPT subjects: a comparison to bvFTD. Neurology. 2011;77:866–74.

[60] Ferguson HJ, Brunsdon VEA, Bradford EEF. The developmental trajectories of executive function from adolescence to old age. Sci Rep. 2021;11:1382.

[61] Salthouse T. Consequences of age-related cognitive declines. Annu Rev Psychol. 2012;63:201–26.

[62] Poos JM, MacDougall A, van den Berg E, Jiskoot LC, Papma JM, van der Ende EL, et al. Longitudinal Cognitive Changes in Genetic Frontotemporal Dementia Within the GENFI Cohort. Neurology. 2022;99:e281–e95.

[63] Adnan A, Beaty R, Lam J, Spreng RN, Turner GR. Intrinsic default-executive coupling of the creative aging brain. Soc Cogn Affect Neurosci. 2019;14:291–303.

[64] Kupis L, Goodman ZT, Kornfeld S, Hoang S, Romero C, Dirks B, et al. Brain Dynamics Underlying Cognitive Flexibility Across the Lifespan. Cereb Cortex. 2021;31:5263–74.

[65] Rohrer JD. Structural brain imaging in frontotemporal dementia. Biochim Biophys Acta. 2012;1822:325–32.

[66] Borroni B, Alberici A, Cercignani M, Premi E, Serra L, Cerini C, et al. Granulin mutation drives brain damage and reorganization from preclinical to symptomatic FTLD. Neurobiol Aging. 2012;33:2506–20.

[67] Rohrer JD, Warren JD, Modat M, Ridgway GR, Douiri A, Rossor MN, et al. Patterns of cortical thinning in the language variants of frontotemporal lobar degeneration. Neurology. 2009;72:1562–9.

[68] Ramanan S, Halai AD, Garcia-Penton L, Perry AG, Patel N, Peterson KA, et al. The neural substrates of transdiagnostic cognitive-linguistic heterogeneity in primary progressive aphasia. Alzheimers Res Ther. 2023;15:219.

[69] Mesulam MM. From sensation to cognition. Brain. 1998;121:1013–52.

[70] Friedman NP, Robbins TW. The role of prefrontal cortex in cognitive control and executive function. Neuropsychopharmacology. 2022;47:72–89.

[71] Murley AG, Rowe JB. Neurotransmitter deficits from frontotemporal lobar degeneration. Brain. 2018;141:1263–85.

[72] Hedden T, Van Dijk KR, Becker JA, Mehta A, Sperling RA, Johnson KA, Buckner RL. Disruption of functional connectivity in clinically normal older adults harboring amyloid burden. J Neurosci. 2009;29:12686–94.

[73] Gabrielyan M, Tisdall MD, Kammer C, Higgins C, Arratia PE, Detre JA. A perfusion phantom for ASL MRI based on impinging jets. Magnetic Resonance in Medicine. 2021;86:1145–58.

[74] Adebimpe A, Bertolero M, Dolui S, Cieslak M, Murtha K, Baller EB, et al. ASLPrep: a platform for processing of arterial spin labeled MRI and quantification of regional brain perfusion. Nat Methods. 2022;19:683–6.

[75] Tsvetanov KA, Ye Z, Hughes L, Samu D, Treder MS, Wolpe N, et al. Activity and Connectivity Differences Underlying Inhibitory Control Across the Adult Life Span. J Neurosci. 2018;38:7887–900.

