## Supplementary for "Frontoparietal network integrity supports cognitive function despite atrophy and hypoperfusion in pre-symptomatic frontotemporal dementia: multimodal analysis of brain function, structure and perfusion"

**Supplementary materials**

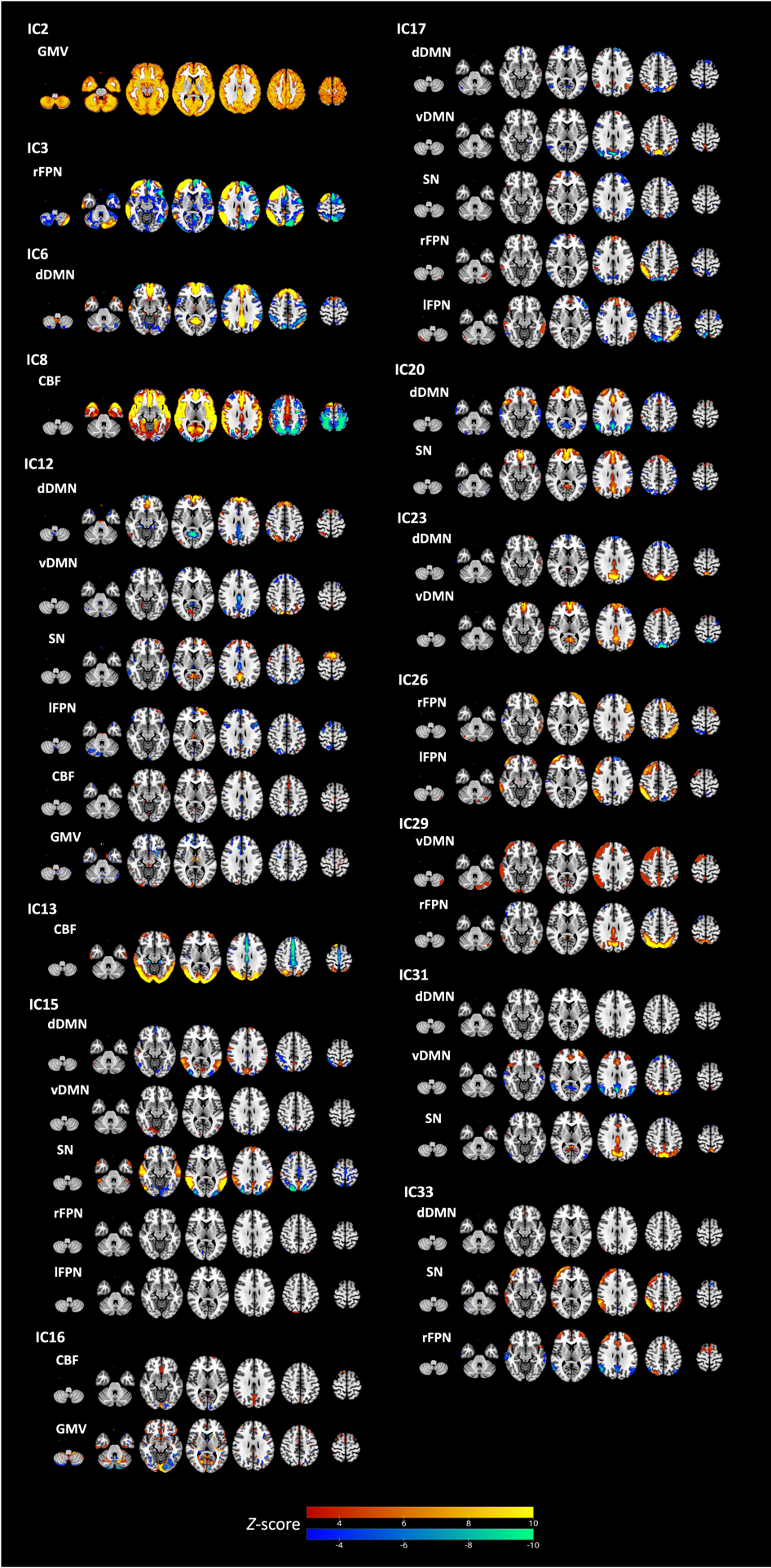

**Supplementary Figure 1.** Brain visualization of linked independent component analysis output components of interest (not shown in the main manuscript). For visualization the spatial map threshold is set to 3 < |*Z*| < 10. Neuroimaging modalities with significant values beyond the threshold are shown. Abbreviations: CBF, cerebral blood flow; dDMN, dorsal default mode network; vDMN, ventral default mode network; rFPN, right frontoparietal network; lFPN, left frontoparietal network; GMV, grey matter volume; SN, salience network.

**Supplementary Figure 2**. Component 1 of PCA of tests examining executive functions only.

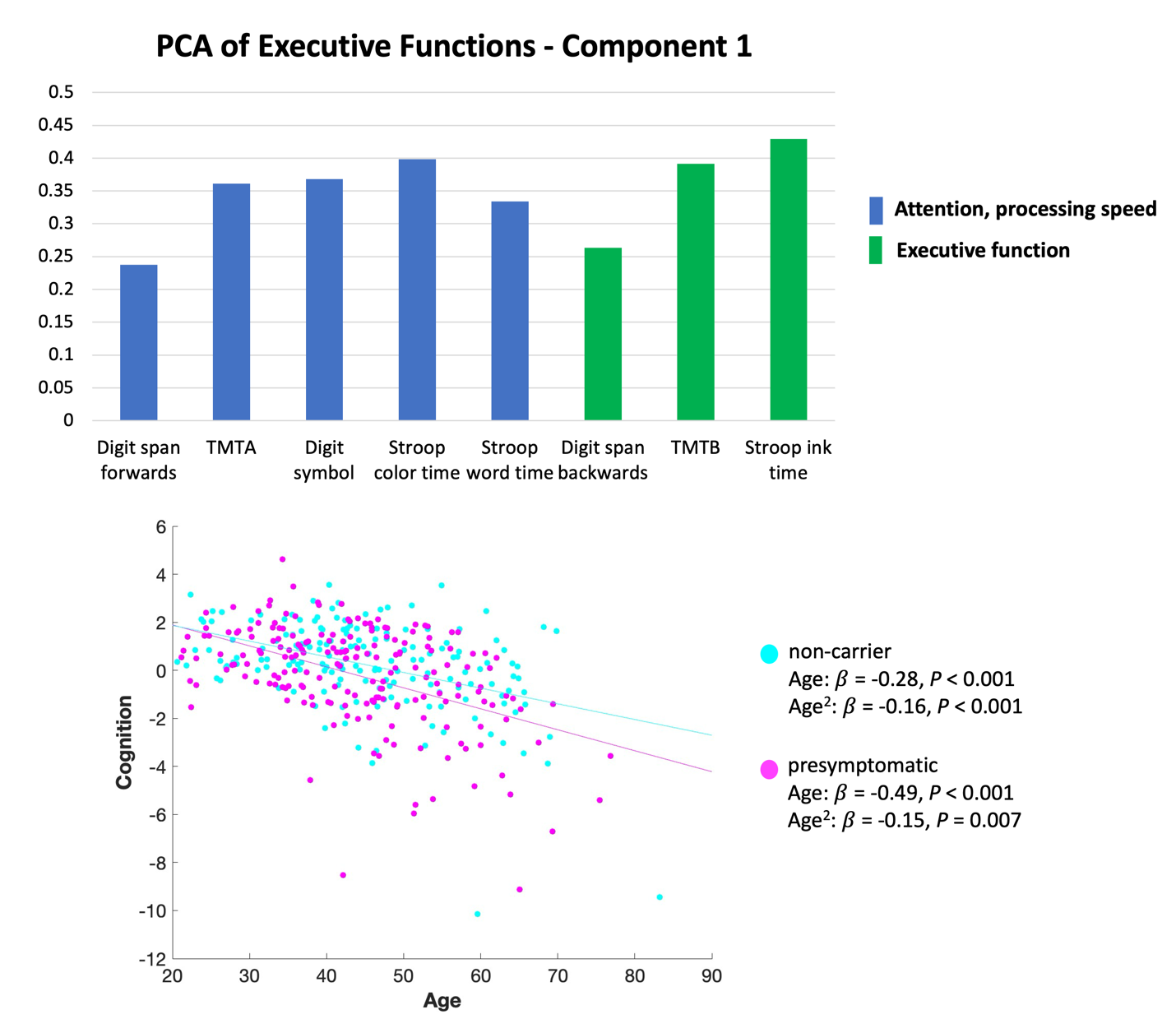

**Supplementary Table 1.** Multiple regression results of the linked independent component analysis components of interest (IC) predicting cognition component 1 (global cognition).

| **Cognition PCA component 1 ~ IC*Group*Age^2 + Gender + Total brain volume + Site** | | | | | | | | | | | | | | | |
| --- | --- | --- | --- | --- | --- | --- | --- | --- | --- | --- | --- | --- | --- | --- | --- |
|  | | **IC** | | **Group** | | **Age** | | **Age^2** | | **IC:Group** | | **IC:Group:Age** | | **IC:Group:Age^2** | |
| IC | Variance explained | β | *P* | β | *P* | β | *P* | β | *P* | β | *P* | β | *P* | β | *P* |
| 1 | 14.23% | 0.08 | 0.19 | -0.08 | 0.18 | -0.37 | **<0.001** | -0.16 | **<0.001** | 0.08 | 0.17 | 0.05 | 0.32 | -0.02 | 0.57 |
| 2 | 9.81% | -0.08 | 0.32 | -0.07 | 0.26 | -0.42 | **<0.001** | -0.14 | **<0.001** | -0.10 | 0.10 | 0.08 | 0.11 | 0.04 | 0.28 |
| 3 | 6.56% | 0.01 | 0.83 | -0.08 | 0.17 | -0.42 | **<0.001** | -0.14 | **<0.001** | 0.01 | 0.91 | -0.05 | 0.34 | 0.07 | 0.07 |
| 4 | 5.10% | -0.06 | 0.33 | -0.08 | 0.17 | -0.40 | **<0.001** | -0.14 | **<0.001** | 0.003 | 0.96 | -0.05 | 0.27 | 0.01 | 0.82 |
| 5 | 4.09% | 0.02 | 0.75 | -0.09 | 0.13 | -0.32 | **<0.001** | -0.14 | **<0.001** | -0.02 | 0.70 | 0.06 | 0.26 | 0.06 | 0.13 |
| 6 | 3.42% | -0.07 | 0.24 | -0.07 | 0.24 | -0.40 | **<0.001** | -0.15 | **<0.001** | <0.001 | 0.99 | -0.02 | 0.59 | 0.003 | 0.92 |
| 7 | 2.18% | -0.04 | 0.53 | -0.08 | 0.17 | -0.38 | **<0.001** | -0.14 | **<0.001** | 0.01 | 0.91 | 0.05 | 0.37 | 0.02 | 0.57 |
| 8 | 1.91% | 0.03 | 0.64 | -0.07 | 0.22 | -0.41 | **<0.001** | -0.14 | **0.001** | -0.13 | 0.03 | 0.03 | 0.55 | 0.05 | 0.15 |
| 9 | 2.14% | 0.12 | 0.08 | -0.07 | 0.22 | -0.31 | **<0.001** | -0.09 | 0.039 | -0.02 | 0.66 | -0.001 | 0.99 | 0.04 | 0.25 |
| 12 | 2.13% | 0.06 | 0.55 | -0.10 | 0.11 | -0.41 | **<0.001** | -0.14 | **<0.001** | -0.02 | 0.73 | -0.02 | 0.67 | 0.01 | 0.90 |
| 13 | 1.02% | -0.09 | 0.09 | -0.09 | 0.13 | -0.39 | **<0.001** | -0.16 | **<0.001** | 0.08 | 0.15 | -0.04 | 0.42 | -0.01 | 0.85 |
| 15 | 2.63% | -0.09 | 0.13 | -0.08 | 0.16 | -0.41 | **<0.001** | -0.14 | **<0.001** | -0.03 | 0.63 | 0.03 | 0.51 | 0.02 | 0.66 |
| 16 | 1.14% | -0.09 | 0.18 | -0.08 | 0.19 | -0.37 | **<0.001** | -0.13 | **0.001** | -0.08 | 0.15 | 0.03 | 0.47 | 0.05 | 0.18 |
| 17 | 2.38% | 0.02 | 0.73 | -0.09 | 0.13 | -0.41 | **<0.001** | -0.15 | **<0.001** | 0.01 | 0.84 | 0.01 | 0.82 | -0.01 | 0.82 |
| 20 | 1.81% | -0.01 | 0.86 | -0.08 | 0.18 | -0.40 | **<0.001** | -0.15 | **<0.001** | -0.01 | 0.92 | 0.06 | 0.16 | 0.05 | 0.12 |
| 23 | 1.47% | 0.09 | 0.15 | -0.09 | 0.14 | -0.41 | **<0.001** | -0.15 | **<0.001** | 0.04 | 0.55 | -0.03 | 0.54 | -0.01 | 0.84 |
| 26 | 1.59% | -0.003 | 0.96 | -0.07 | 0.22 | -0.41 | **<0.001** | -0.16 | **<0.001** | 0.08 | 0.14 | -0.07 | 0.19 | -0.06 | 0.14 |
| 29 | 1.45% | 0.05 | 0.41 | -0.10 | 0.10 | -0.43 | **<0.001** | -0.13 | **0.001** | 0.06 | 0.30 | 0.06 | 0.24 | -0.04 | 0.26 |
| 31 | 1.22% | 0.03 | 0.56 | -0.09 | 0.13 | -0.41 | **<0.001** | -0.15 | **<0.001** | 0.03 | 0.61 | -0.02 | 0.71 | <0.001 | 0.99 |
| 33 | 1.17% | -0.02 | 0.73 | -0.09 | 0.12 | -0.43 | **<0.001** | -0.17 | **<0.001** | 0.11 | 0.05 | 0.01 | 0.91 | -0.08 | 0.03 |

P values in bold are statistically significant

**Supplementary Table 2.** Multiple regression results of the linked independent component analysis components of interest (IC) predicting cognition component 2 (executive functions).

| **Cognition PCA component 2 ~ IC*Group*Age^2 + Gender + Total brain volume + Site** | | | | | | | | | | | | | | | |
| --- | --- | --- | --- | --- | --- | --- | --- | --- | --- | --- | --- | --- | --- | --- | --- |
|  | | **IC** | | **Group** | | **Age** | | **Age^2** | | **IC:Group** | | **IC:Group:Age** | | **IC:Group:Age^2** | |
| IC | Variance explained | β | *P* | β | *P* | β | *P* | β | *P* | β | *P* | β | *P* | β | *P* |
| 1 | 14.23% | 0.04 | 0.57 | -0.02 | 0.79 | 0.07 | 0.30 | -0.02 | 0.60 | 0.12 | 0.08 | 0.09 | 0.13 | -0.03 | 0.50 |
| 2 | 9.81% | -0.11 | 0.21 | -0.002 | 0.97 | -0.02 | 0.70 | -0.07 | 0.10 | -0.19 | **0.007** | -0.02 | 0.72 | 0.03 | 0.44 |
| 3 | 6.56% | 0.01 | 0.93 | -0.03 | 0.63 | -0.01 | 0.83 | -0.04 | 0.28 | 0.10 | 0.12 | -0.01 | 0.86 | -0.05 | 0.31 |
| 4 | 5.10% | 0.06 | 0.40 | -0.01 | 0.90 | -0.01 | 0.83 | -0.04 | 0.34 | 0.07 | 0.26 | 0.06 | 0.26 | -0.09 | **0.029** |
| 5 | 4.09% | -0.01 | 0.83 | 0.005 | 0.94 | -0.05 | 0.44 | -0.03 | 0.42 | 0.10 | 0.14 | 0.11 | 0.06 | -0.11 | **0.007** |
| 6 | 3.42% | -0.07 | 0.32 | -0.03 | 0.67 | -0.01 | 0.86 | -0.02 | 0.63 | 0.05 | 0.48 | -0.04 | 0.44 | -0.06 | 0.065 |
| 7 | 2.18% | -0.05 | 0.47 | -0.0002 | 0.99 | -0.01 | 0.84 | -0.03 | 0.40 | 0.16 | **0.01** | -0.01 | 0.91 | -0.15 | **0.002** |
| 8 | 1.91% | 0.07 | 0.25 | -0.02 | 0.80 | -0.01 | 0.90 | -0.02 | 0.66 | < 0.001 | 0.99 | 0.02 | 0.71 | -0.01 | 0.77 |
| 9 | 2.14% | -0.02 | 0.82 | -0.01 | 0.86 | 0.02 | 0.80 | -0.01 | 0.76 | 0.01 | 0.93 | 0.06 | 0.34 | -0.03 | 0.56 |
| 12 | 2.13% | 0.13 | 0.28 | -0.01 | 0.83 | < 0.001 | 0.99 | -0.03 | 0.49 | 0.03 | 0.67 | 0.08 | 0.16 | 0.06 | 0.24 |
| 13 | 1.02% | -0.01 | 0.91 | -0.03 | 0.67 | < 0.001 | 0.99 | -0.04 | 0.39 | 0.18 | **0.004** | 0.06 | 0.33 | -0.07 | 0.11 |
| 15 | 2.63% | 0.24 | < 0.001 | -0.03 | 0.59 | 0.02 | 0.71 | -0.03 | 0.41 | 0.02 | 0.76 | -0.01 | 0.81 | 0.01 | 0.83 |
| 16 | 1.14% | 0.11 | 0.11 | -0.02 | 0.80 | -0.002 | 0.97 | -0.03 | 0.41 | -0.09 | 0.17 | -0.06 | 0.30 | 0.04 | 0.39 |
| 17 | 2.38% | -0.14 | 0.05 | -0.03 | 0.67 | 0.01 | 0.89 | -0.02 | 0.67 | 0.04 | 0.52 | 0.13 | **0.02** | 0.04 | 0.45 |
| 20 | 1.81% | -0.02 | 0.80 | -0.03 | 0.70 | -0.03 | 0.66 | -0.05 | 0.21 | 0.02 | 0.74 | 0.04 | 0.41 | -0.08 | **0.04** |
| 23 | 1.47% | -0.08 | 0.26 | -0.02 | 0.72 | -0.01 | 0.86 | -0.03 | 0.47 | 0.003 | 0.97 | -0.02 | 0.71 | -0.01 | 0.76 |
| 26 | 1.59% | -0.03 | 0.61 | -0.03 | 0.60 | -0.03 | 0.65 | -0.04 | 0.36 | -0.03 | 0.64 | 0.03 | 0.65 | 0.10 | 0.05 |
| 29 | 1.45% | -0.05 | 0.43 | 0.00 | 0.99 | -0.02 | 0.72 | -0.09 | **0.04** | -0.05 | 0.48 | -0.04 | 0.42 | 0.07 | 0.11 |
| 31 | 1.22% | -0.09 | 0.18 | -0.04 | 0.52 | 0.01 | 0.89 | -0.01 | 0.74 | -0.06 | 0.38 | 0.06 | 0.32 | 0.08 | 0.07 |
| 33 | 1.17% | -0.15 | 0.02 | -0.06 | 0.36 | 0.001 | 0.99 | -0.03 | 0.54 | 0.05 | 0.40 | 0.03 | 0.63 | 0.06 | 0.12 |

P values in bold are statistically significant
